# Assessing the feasibility and impact of clinical trial trustworthiness checks via an application to Cochrane Reviews: Stage 2 of the INSPECT-SR project

**DOI:** 10.1101/2024.11.25.24316905

**Authors:** Jack Wilkinson, Calvin Heal, Georgios A Antoniou, Ella Flemyng, Love Ahnström, Alessandra Alteri, Alison Avenell, Timothy Hugh Barker, David N Borg, Nicholas JL Brown, Rob Buhmann, Jose A Calvache, Rickard Carlsson, Lesley-Anne Carter, Aidan G Cashin, Sarah Cotterill, Kenneth Färnqvist, Michael C Ferraro, Steph Grohmann, Lyle C Gurrin, Jill A Hayden, Kylie E Hunter, Natalie Hyltse, Lukas Jung, Ashma Krishan, Silvy Laporte, Toby J Lasserson, David RT Laursen, Sarah Lensen, Wentao Li, Tianjing Li, Jianping Liu, Clara Locher, Zewen Lu, Andreas Lundh, Antonia Marsden, Gideon Meyerowitz-Katz, Ben W Mol, Zachary Munn, Florian Naudet, David Nunan, Neil E O’Connell, Natasha Olsson, Lisa Parker, Eleftheria Patetsini, Barbara Redman, Sarah Rhodes, Rachel Richardson, Martin Ringsten, Ewelina Rogozińska, Anna Lene Seidler, Kyle Sheldrick, Katie Stocking, Emma Sydenham, Hugh Thomas, Sofia Tsokani, Constant Vinatier, Colby J Vorland, Rui Wang, Bassel H Al Wattar, Florencia Weber, Stephanie Weibel, Madelon van Wely, Chang Xu, Lisa Bero, Jamie J Kirkham

**Author notes:** Joint first authorship (Jack Wilkinson, Calvin Heal). Joint senior authorship (Lisa Bero, Jamie J Kirkham).

## Abstract

**Background:** The aim of the INSPECT-SR project is to develop a tool to identify problematic RCTs in systematic reviews. In Stage 1 of the project, a list of potential trustworthiness checks was created. The checks on this list must be evaluated to determine which should be included in the INSPECT-SR tool.

**Methods:** We attempted to apply 72 trustworthiness checks to RCTs in 50 Cochrane Reviews. For each, we recorded whether the check was passed, failed or possibly failed, or whether it was not feasible to complete the check. Following application of the checks, we recorded whether we had concerns about the authenticity of each RCT. We repeated each meta-analysis after removing RCTs flagged by each check, and again after removing RCTs where we had concerns about authenticity, to estimate the impact of trustworthiness assessment. Trustworthiness assessments were compared to Risk of Bias and GRADE assessments in the reviews.

**Results:** 95 RCTs were assessed. Following application of the checks, assessors had some or serious concerns about the authenticity of 25% and 6% of the RCTs, respectively. Removing RCTs with either some or serious concerns resulted in 22% of meta-analyses having no remaining RCTs. However, many checks proved difficult to understand or implement, which may have led to unwarranted scepticism in some instances. Furthermore, we restricted assessment to meta-analyses with no more than 5 RCTs, which will distort the impact on results. No relationship was identified between trustworthiness assessment and Risk of Bias or GRADE.

**Conclusions:** This study supports the case for routine trustworthiness assessment in systematic reviews, as problematic studies do not appear to be flagged by Risk of Bias assessment. The study produced evidence on the feasibility and impact of trustworthiness checks. These results will be used, in conjunction with those from a subsequent Delphi process, to determine which checks should be included in the INSPECT-SR tool.

**Plain language summary:** Systematic reviews collate evidence from randomised controlled trials (RCTs) to find out whether health interventions are safe and effective. However, it is now recognised that the findings of some RCTs are not genuine, and some of these studies appear to have been fabricated. Various checks for these “problematic” RCTs have been proposed, but it is necessary to evaluate these checks to find out which are useful and which are feasible. We applied a comprehensive list of “trustworthiness checks” to 95 RCTs in 50 systematic reviews to learn more about them, and to see how often performing the checks would lead us to classify RCTs as being potentially inauthentic. We found that applying the checks led to concerns about the authenticity of around 1 in 3 RCTs. However, we found that many of the checks were difficult to perform and could have been misinterpreted. This might have led us to be overly sceptical in some cases. The findings from this study will be used, alongside other evidence, to decide which of these checks should be performed routinely to try to identify problematic RCTs, to stop them from being mistaken for genuine studies and potentially being used to inform healthcare decisions.

**What is new:**

- An extensive list of potential checks for assessing study trustworthiness was assessed via an application to 95 randomised controlled trials (RCTs) in 50 Cochrane Reviews.
- Following application of the checks, assessors had concerns about the authenticity of 32% of the RCTs.
- If these RCTs were excluded, 22% of meta-analyses would have no remaining RCTs.
- However, the study showed that some checks were frequently infeasible, and others could be easily misunderstood or misinterpreted.
- The study restricted assessment to meta-analyses including five or fewer RCTs, which might distort the impact of applying the checks.

## Background

Systematic reviews of randomised controlled trials (RCTs) aim to include all trials that address the review question and meet the prespecified eligibility criteria. There is an understanding that RCTs included in a systematic review should be scrutinised for their internal validity, for example, using Risk of Bias tools (1, 2). These assessments require that the reviewer can trust what is written in a trial report to be an authentic account of what took place. However, this no longer appears to be tenable as a default assumption, as recent large-scale assessments have cast doubt on the veracity of many RCTs submitted to journals (3) or published in systematic reviews (4). Recent examples, such as ivermectin for COVID-19, illustrate how the failure to routinely interrogate the authenticity of eligible RCTs in systematic reviews allows fake studies to influence patient care (5).

Cochrane defines ‘*problematic studies’* as studies where there are ‘serious questions about the trustworthiness of the data or findings’ (6). Problematic studies could represent instances of academic misconduct such as research fraud, or could be the result of critical errors in trial processes. Cochrane policy, introduced in 2021, states that potentially problematic RCTs should not be included in a systematic review (6, 7). This prompts the question of what criteria could be used to identify problematic studies, which may appear to be high-quality on the basis of traditional Risk of Bias assessment (8). Cochrane’s implementation guidance recognises that a number of methods for identifying problematic studies have been proposed, but does not recommend a method at this time.

The aim of the INSPECT-SR (INveStigating ProblEmatic Clinical Trials in Systematic Reviews) project is to develop a tool that can be used by systematic reviewers to assess the trustworthiness of RCTs (9). Several tools have recently been proposed for this purpose (10–14). However, none of these have involved a comprehensive evaluation and subsequent selection of potential trustworthiness checks. In Stage 1 of the development process, we identified an extensive list of potential trustworthiness checks (15). A tool including all of these checks would not be practicable, and we anticipate that many of the checks will turn out to be infeasible or otherwise not useful. In Stages 2 (application to Cochrane Reviews) and 3 (Delphi survey), the checks on this list will be evaluated to determine which should be included in the final tool. These results will then feed into a series of consensus meetings (Stage 4) which will be used to develop a draft version of the INSPECT-SR tool. The draft tool will then be tested in the assessment of RCTs (Stage 5). Feedback from Stage 5 will be used to finalise the tool. The current study describes Stage 2 of the project, in which the identified checks were applied to RCTs included in a sample of Cochrane Reviews, in order to evaluate their feasibility and impact on review results, and to evaluate how often assessors had concerns about the authenticity of RCTs after applying the checks.

## Methods

A protocol describing the INSPECT-SR project methods has previously been published (9). We undertook a large, collaborative project in which assessors applied a series of 72 trustworthiness checks to RCTs included in 50 Cochrane Reviews. The University of Manchester Ethics tool was used to determine that ethical approval was not required for this study (30th Sept 2022).

### Description of trustworthiness checks

Prior to this exercise, a list of trustworthiness checks was assembled using a scoping review (16), qualitative study (17) and survey of experts (15). This list contained 116 checks arranged into five domains: *Inspecting results in the paper, Inspecting the research team and their work, Inspecting conduct, governance and transparency, Inspecting text and publication details,* and *Inspecting individual participant data*. In the current study we only considered the first four domains, as individual participant data are not generally available during systematic reviews and meta-analyses based on aggregate data; nor were they available to us. An extension to the INSPECT-SR tool based on the checks in the fifth domain, which can be applied when individual participant data are available, ‘INSPECT-IPD’, has been funded for development (Reference: NIHR30355). The first four domains included 76 checks (Tables 1 and 2). We made some modifications to the list in preparation for the current study, in consultation with the project expert advisory panel. This included refining the language of some items to improve clarity. To assist assessors in applying the checks, we drafted brief explanations for each check (S Tables 1 to 4). Four checks (checks 45, 66, 67, 72, Tables 1 and 2) were not assessed as they were not considered practicable in the context of the present study. Consequently, 72 checks were assessed.

**Table 1:**
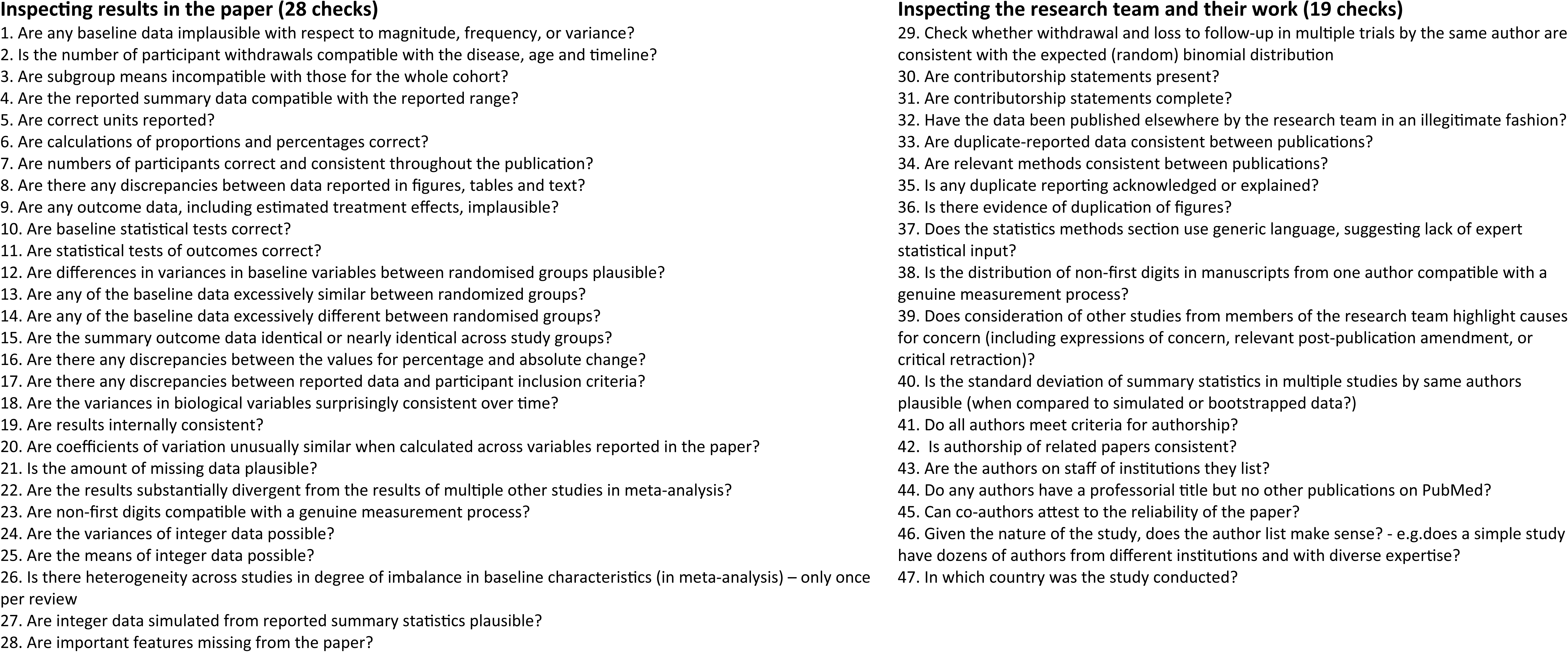
Trustworthiness checks in the first and second domains of the assessed list.

**Table 2:**
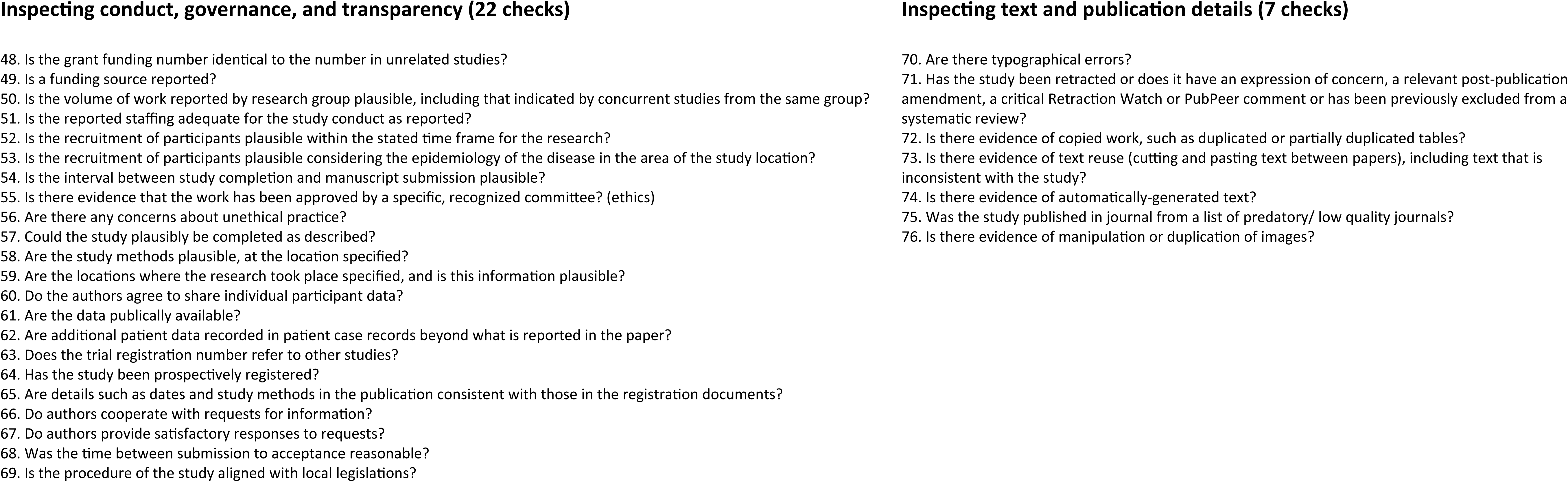
Trustworthiness checks in the third and fourth domains of the assessed list.

### Description of assessors

The INSPECT-SR working group includes a core management group and an expert advisory panel. Members of both were invited to act as assessors for the current study. We also invited additional collaborators who had expressed an interest in contributing to the development process. Collaborators were identified from a variety of sources. We invited attendees at presentations relating to the project to contact us to express an interest, and also invited individuals who had expressed an interest in the topic to JW using personalised emails and via social media. All assessors were considered to have sufficient expertise in research methods (specifically, to evaluate RCTs) to enable them to undertake the assessment. We did not require assessors to hold any particular qualification however. We did not require assessors to have specialist expertise relating to research integrity (for example, use of forensic statistical methods or investigation of misconduct cases), as a key objective was to learn about the usefulness and feasibility of the checks when applied by potential users of the INSPECT-SR tool (i.e. systematic reviewers, researchers, peer reviewers) who would not be expected to possess this specialist knowledge. Assessors who were considered to have made a substantial contribution to data acquisition and critical review of manuscript drafts, were given the option to co-author the manuscript.

### Selection of Cochrane Reviews and RCTs

The sample size of 50 Cochrane reviews represented a number that was considered feasible to complete, while facilitating the evaluation of feasibility and impact of applying the checks across different topic areas. A preliminary pilot was conducted on a small number of RCTs to confirm this. The 50 reviews were purposefully selected from the Cochrane Library. To be eligible, a review could not be authored or co-authored by the assessor, and could not contain RCTs authored or co-authored by the assessor, to prevent any conflict of interest in conducting the assessment. As a feasibility requirement, we also required that the review contained at least one (meta-) analysis containing one to five RCTs. For brevity, we use the term ‘meta-analysis’ in this article to describe an analysis which produces a pooled average estimate and confidence interval for a treatment effect on an outcome based on the included studies, recognising that, when there is only one study, this involves reporting the estimate and confidence interval from that study. The RCTs in the first eligible meta-analysis in the review were subjected to the trustworthiness assessment, as a feasibility constraint. We also required that the review had not already undergone a trustworthiness assessment as part of the review process, since this could have resulted in the prior removal of problematic studies, distorting our assessment. Assessors were asked to suggest a topic with which they were broadly familiar. We attempted to match assessors to review topics, to replicate a typical scenario in which INSPECT-SR would be used (a systematic review would often be undertaken by someone with some relevant subject-matter knowledge). We then selected the most recent Cochrane Review relating to the topic suggested by the assessor that met the eligibility criteria. Assessors did not always have subject-matter knowledge relating to the review(s) they assessed, however. For example, some assessors were primarily methodologists, with limited clinical knowledge of the subject matter. For these people, we attempted to select review topics to cover a broad range of health areas. We asked each assessor to record their familiarity with the review topic during data extraction (little or no familiarity, some familiarity, or high familiarity).

### Data extraction and trustworthiness assessment

A bespoke data extraction form was produced, and was revised following piloting on a small number of RCTs, and can be accessed at https://osf.io/9pyw2/. Assessors were informed of software that could be used to implement some of the statistical checks. Examples include the scrutiny package in R (18), online applications created to implement some checks e.g. (19, 20), or Microsoft Excel (21) for basic statistical checks, but it was not a requirement to use any particular software to undertake the assessment. For each Cochrane Review, the assessor extracted data and applied the list of checks to each RCT in the meta-analysis. An exception was check 26 - *Is there heterogeneity across studies in degree of imbalance in baseline characteristics (in meta-analysis)*, which was assessed only once per review. The assessor extracted the year of publication for each RCT, the summary data entered in the meta-analysis, and Risk of Bias and GRADE (Grading of Recommendations Assessment, Development and Evaluation) (22) assessments as presented in the Cochrane Review.

A second assessor checked the accuracy and completeness of this information following extraction. Any disagreements were resolved by discussion between assessors and a third team member (JW). The assessor attempted to apply each of the 72 checks to the trial, selecting one of four response options: not feasible; passed the check; possible fail; fail. For each check, assessors were asked to supply free text to explain their assessment. The country or countries in which the RCT was conducted was also recorded. After applying the checks, assessors recorded their answer to the question ‘’Do you have concerns about the authenticity of this study?” using one of four response options: no; some concerns; serious concerns; don’t know. Assessors were asked if they had performed any additional checks (not included on the list) and if so, to describe both the checks and the results of applying them. There was space for the assessor to add any additional information, and to provide an estimate of how many hours it took them to assess the RCT. The intention had been for one assessor to assess all of the RCTs in the review, before checking by a second assessor. However, some assessors failed to complete the assessment of all RCTs in their allocated review, and so for several reviews the RCTs were split between two assessors, before being checked by a third assessor.

### Statistical analysis

We summarised trial and Cochrane Review characteristics, and the responses for each check. We calculated how often assessors had concerns about study authenticity. We evaluated the impact of applying each check by comparing the analysis/ meta-analysis including all trials as per the review to a version in which any RCTs flagged by the check were removed, in terms of the numbers of trials, sample size, change in effect estimate, 95% confidence interval width, heterogeneity, and change in inference.

The first two of these metrics were assessed over all reviews, while the remainder were assessed separately for binary and continuous outcomes. We used the metafor package (23) in R to perform all meta-analyses, using odds ratios to summarise treatment effects with binary outcomes, and standardised mean differences to summarise treatment effects with continuous outcomes. Random effects meta-analyses using the DerSimonian and Laird (24) method were performed, as the most typical method employed in systematic reviews (25, 26).

We assessed potential redundancies among the checks by plotting the responses for each check for each RCT in an array. We made the post-hoc decision to undertake a hierarchical cluster analysis, using complete agglomeration based on Gower dissimilarity, as implemented in the cluster package in R (27). We used multinomial regression to assess the relationship between trustworthiness assessment and each Risk of Bias domain, and ordinal regression (proportional odds logistic regression) to consider the relationship between the GRADE assessment and the number of trials flagged for concerns. We used likelihood ratio tests for inference following regression model fits. We conducted an additional analysis which had not been specified in the protocol, where we evaluated the relationship between the assessment for each check and the overall assessment of the trial using the N-1 chi-squared test (28), to determine which checks were influential in reaching an overall assessment. The N-1 chi-squared test was used in anticipation of small expected counts (29). This analysis was performed in trials where the check was considered to be feasible, and the assessments were analysed as ‘passed’ vs ‘fail or possible fail’. We used a post-hoc significance threshold of 1% to highlight checks associated with the overall assessment, creating contingency tables (outcome of check vs overall assessment) for these checks to determine whether failing the check was associated with an assessor having overall concerns. We categorised the free-text responses to the question asking how long it took to complete the assessment in a post-hoc fashion (less than 90 minutes, 90 minutes to 3 hours, more than 3 hours). The dataset and analysis code for this study are available at https://osf.io/9pyw2/.

## Results

We included a total of 95 RCTs from 50 Cochrane Reviews. The reviews were from 24 different Cochrane Groups (S Table 5). Assessors considered themselves to have high familiarity with the review topic for 7/50 (14%) reviews, some familiarity for 20/50 (40%) of reviews, and little or no familiarity for 23/50 (46%). The characteristics of included Cochrane Reviews are shown in Table 3. The median (IQR) number of participants in the assessed RCTs was 71 (40 to 174). 15/95 (16%) were conducted in multiple countries, with the remaining 80 taking place in one of 21 different countries (S Table 6). Twenty-four (26%) RCTs took less than 90 minutes to assess, 29 (31%) took between 90 minutes and 3 hours, and 40 (42%) took more than 3 hours.

**Table 3:**
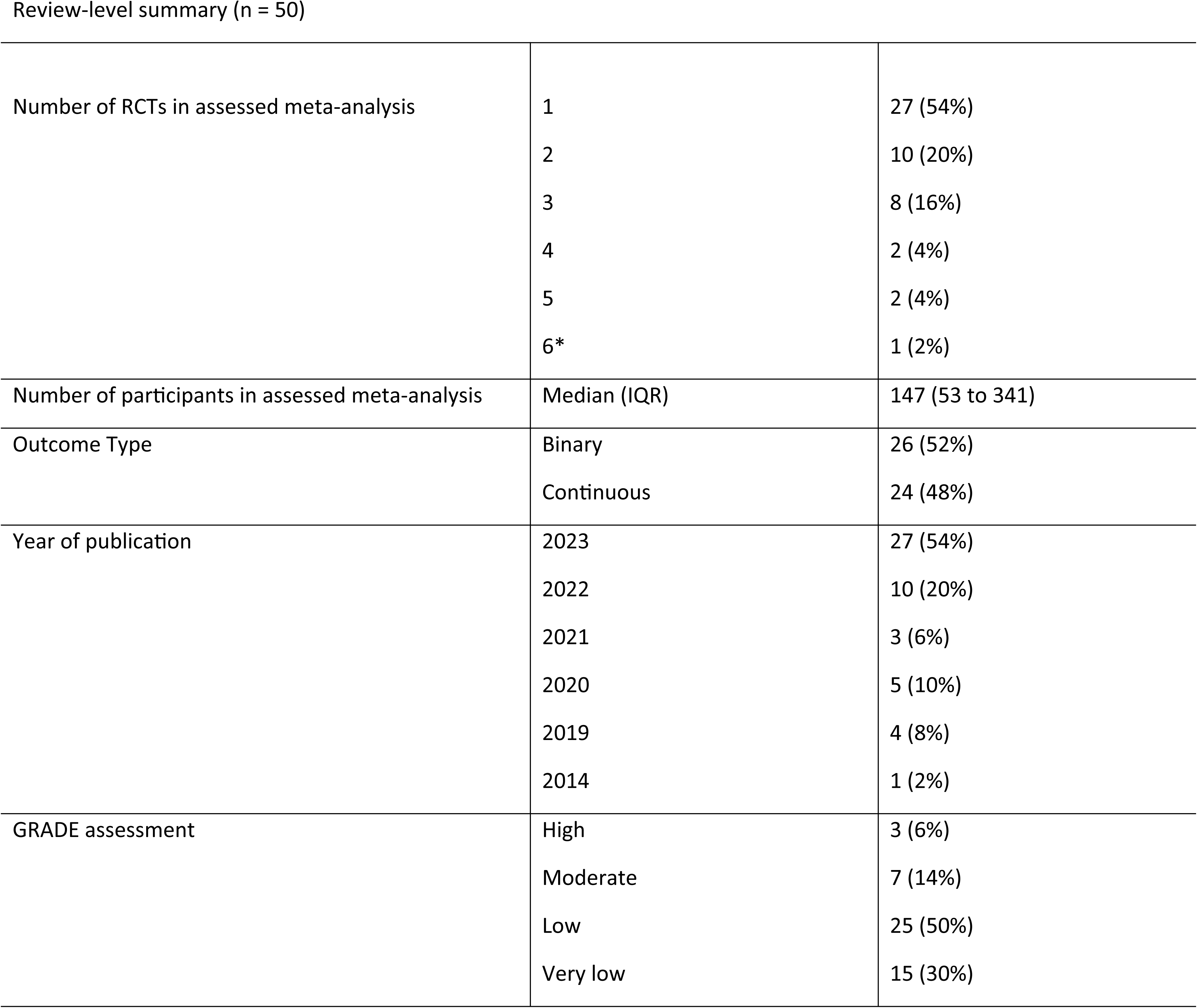
Characteristics for 50 Cochrane Reviews assessed in the study. Frequency (%) or median (1^st^ quartile to 3^rd^ quartile) *Assessed in error, included in analysis.

### Responses to individual trustworthiness checks

Figure 1 and S Table 7 summarise the responses for each check, and S Figure 1 shows the study-level responses for each check. S Figure 2 shows how the checks are clustered in the dataset. Missing data for trustworthiness checks were infrequent, with only one check having missing data for as many as five RCTs (check 42). Check 26 is ‘missing’ for 10 RCTs, as it was only assessed once per review. A number of checks were considered to have ‘failed’ or ‘possibly failed’ often. The five checks most often receiving an assessment of ‘failed’ or ‘possibly failed’ were check 61 - *Are the data publically available?* (81%), check 30 - *Are contributorship statements present?* (69%), check 31 - *Are contributorship statements complete?* (57%), check 64 - *Has the study been prospectively registered?* (56%), check 49 – *Is a funding source reported?* (40%). Some statistical checks frequently resulted in responses of ‘failed’ or ‘possibly failed’. Examples include check 12 - *Are differences in variances in baseline variables between randomised groups plausible?* (28%), check 11 – *Are statistical test [results] of outcomes correct?* (21%).

**Figure 1:**
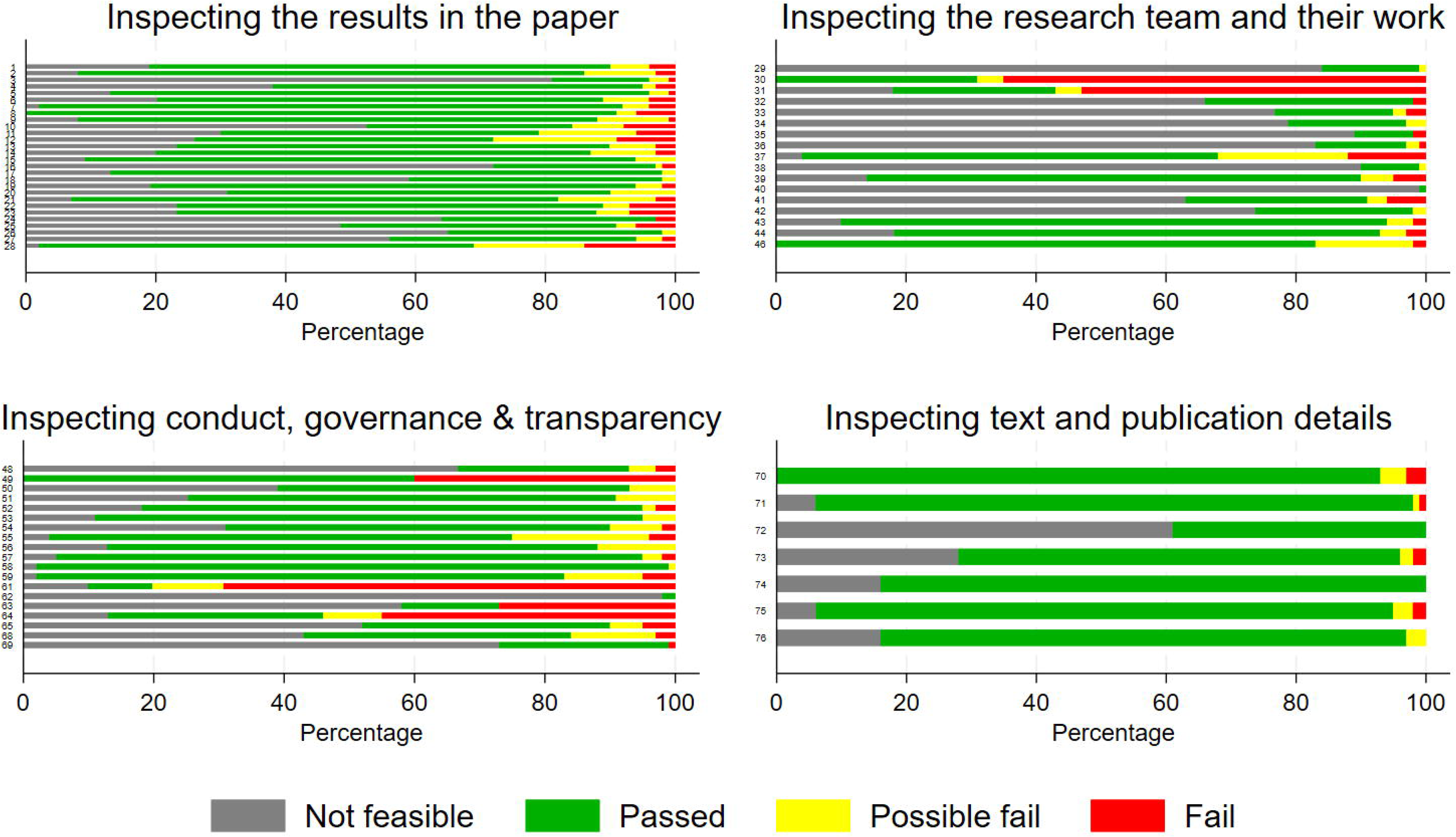
Responses to trustworthiness checks in four domains

**S Figure 1:**
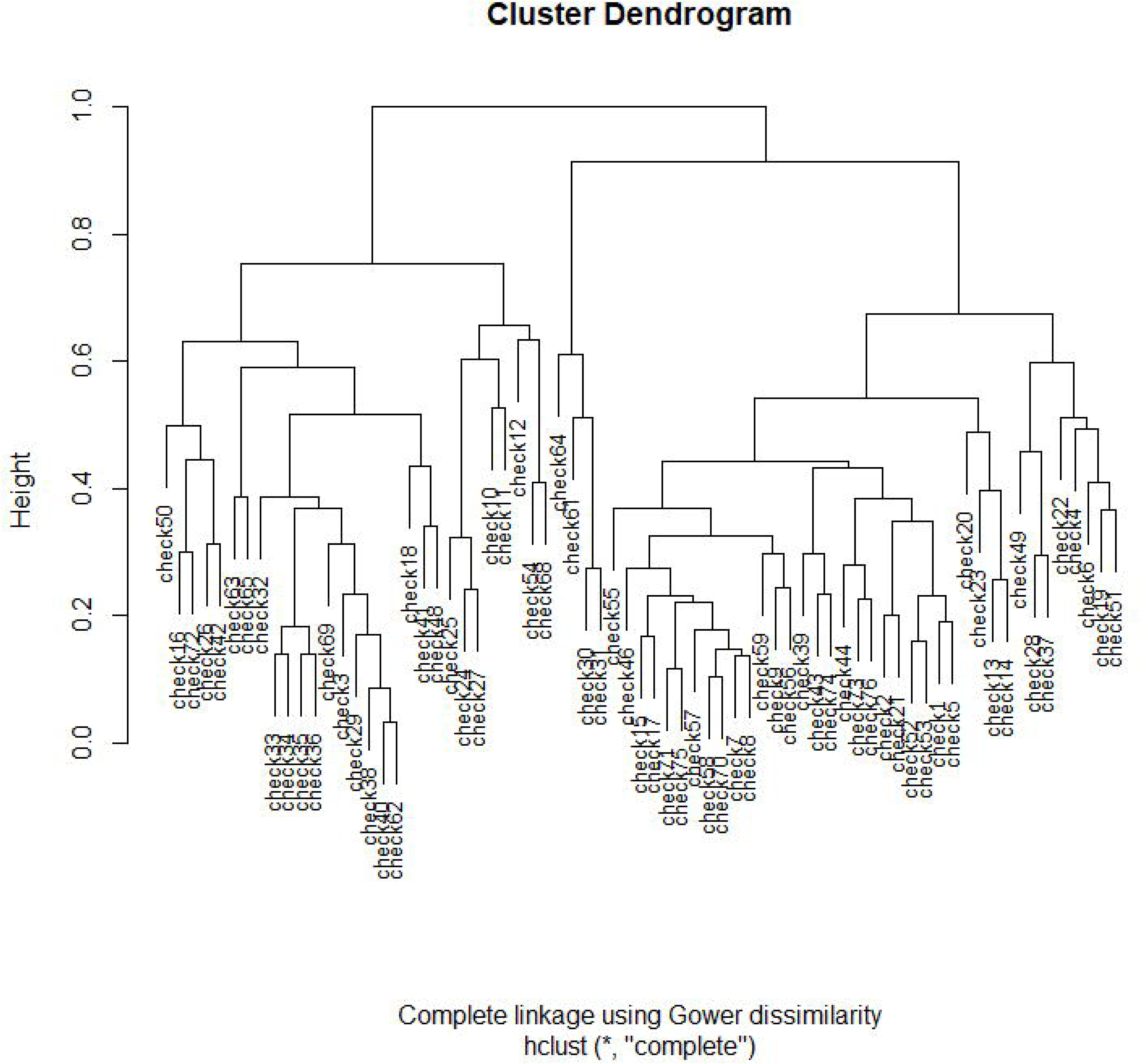
Study-level responses to trustworthiness checks

**S Figure 2:**
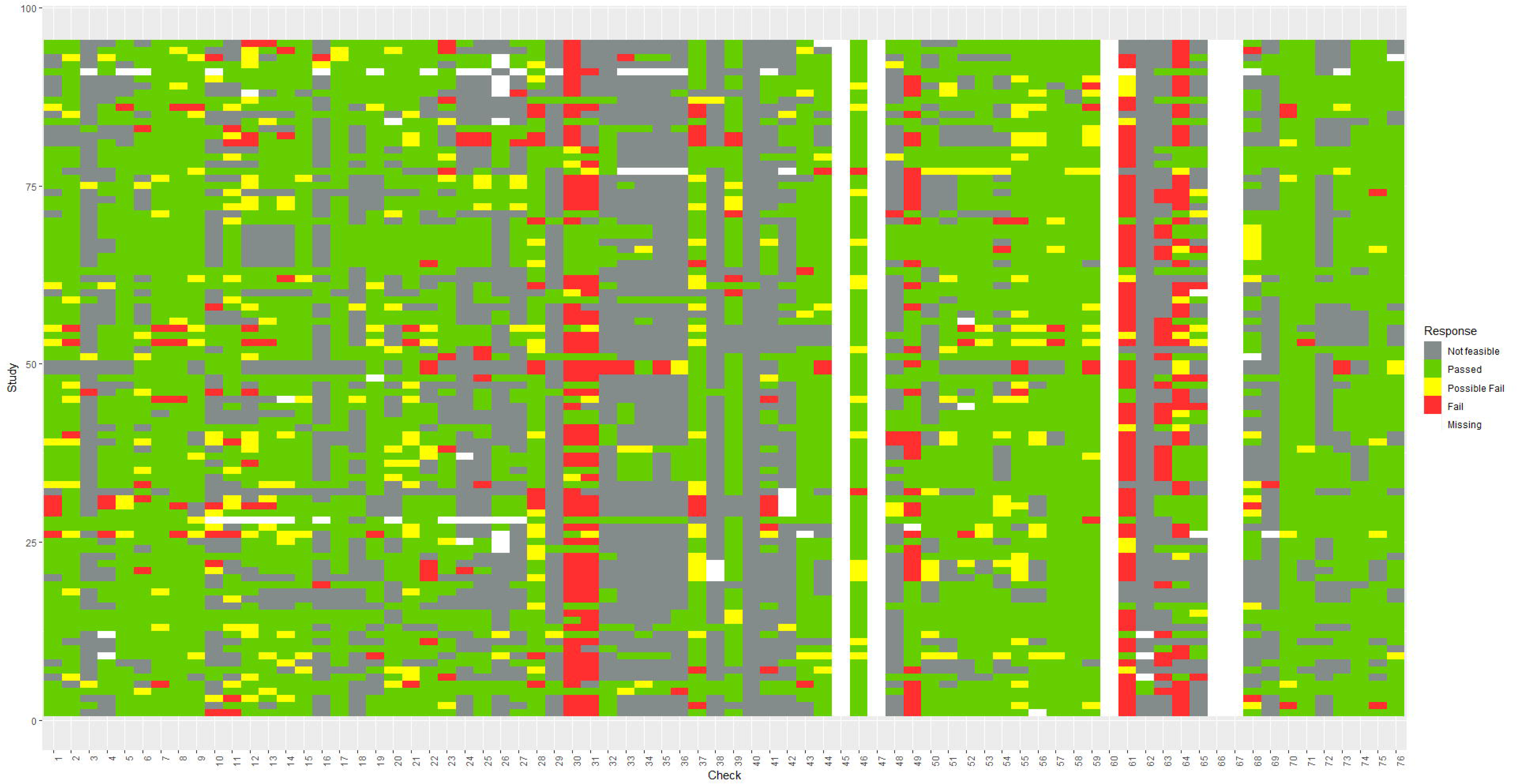
Dendrogram displaying hierarchical clustering of checks using complete agglomeration based on Gower dissimilarity

A number of checks were considered to be infeasible in most cases. The checks most frequently considered infeasible were check 40 - *Is the standard deviation of summary statistics in multiple studies by same authors plausible (when compared to simulated or bootstrapped data?)* (99%), check 62 - *Are additional patient data recorded in patient case records beyond what is reported in the paper? (*98%), check 38 *-Is the distribution of non-first digits in manuscripts from one author compatible with a genuine measurement process?* (90%), check 35 - *Is any duplicate reporting acknowledged or explained?* (89%), and check 29 - *Are withdrawal and loss to follow-up in multiple trials by the same author consistent with the expected (random) binomial distribution?* (84%).

### Overall assessment and relationship to individual checks

Overall, responses to the question “Do you have concerns about the authenticity of this study?” were: no (60/95, 64%); some concerns (24/95, 25%); serious concerns (6/95, 6%). P-values from chi-squared tests looking at the outcome of each check against the overall trustworthiness assessment of the study are shown in S Table 8. Noting that these analyses were post-hoc and exploratory, 19 checks were associated with overall assessment using a 1% significance level. Contingency tables were inspected to examine the nature of these associations (specifically to confirm that failing or possibly failing, rather than passing, a check was associated with the presence of concerns).

Of these 19, there were 11 checks for which failing (as opposed to passing) the check appeared to correlate with an assessment of overall concern: 1. *Are any baseline data implausible with respect to magnitude, frequency, or variance?* (p=0.00001); 2. *Is the number of participant withdrawals compatible with the disease, age and timeline?* (p=0.005); 8. *Are there any discrepancies between data reported in figures, tables and text?* (p = 0.00006), 9. *Are any outcome data, including estimated treatment effects, implausible?* (p = 0.000002), 19. *Are results internally consistent?* (p=0.00008), 37. *Does the statistics methods section use generic language, suggesting lack of expert statistical input?* (p=0.003), 51. *Is the reported staffing adequate for the study conduct as reported?* (p=0.009), 52. *Is the recruitment of participants plausible within the stated time frame for the research?* (p=0.0005), 53. *Is the recruitment of participants plausible considering the epidemiology of the disease in the area of the study location?* (p=0.0004), 56. *Are there any concerns about unethical practice?* (p=0.001), 64. *Has the study been prospectively registered?* (p=0.004).

### Impact of applying the trustworthiness assessments on systematic review results

S Table 9 and S Table 10 show the impact of removing RCTs flagged by each check (considered individually) from meta-analysis, for binary and continuous outcomes respectively. In continuous outcome meta-analyses, removal of RCTs flagged by a check resulted in a median of 4% (IQR 0% to 12.5%, range 0% to 67%) of meta-analyses having no remaining trials. In binary outcome meta-analysis, the corresponding values were 4% (IQR 0% to 8%, range 0% to 73%). The sample size of reviews would be reduced to a median (of means) of 93% (IQR 87% to 97%, range 27% to 100%) of the original size. The median (of means) number of trials that would be removed from meta-analysis was 0.14 (IQR 0.06 to 0.24, range 0 to 1.52).

When RCTs were removed on the basis of the overall assessment (some or serious concerns), 33% of continuous outcome meta-analyses and 12% of binary outcome meta-analyses had no remaining trials. Amongst meta-analyses with at least one RCT remaining, for binary outcome meta-analyses, the mean ROR was 0.98; SE increased by 19% on average; none changed in terms of statistical significance (using a 5% significance threshold); and the mean ratio of confidence interval widths (width expressed as the ratio of upper to lower limit on OR scale) was 4.52. For continuous outcome meta-analyses with at least one RCT remaining, the average change in estimate was-0.02 SDs; SE (and, equivalently, CI width) increased by a mean of 5%; and none of the meta-analyses changed in terms of statistical significance.

### Relationship between trustworthiness assessments, Risk of Bias and GRADE

We only investigated the relationship between overall trustworthiness assessment and risk of bias for reviews using the first version of the Cochrane RoB tool since there were only 10 reviews applying RoB 2. Multinomial regression did not indicate associations between any risk of bias domain and overall concern, with the exception of allocation concealment. However, this was not in the expected direction, with concerns expressed more often for studies with unclear or low bias assessment compared to high bias assessment (p=0.01). The estimated relationship between number of trials flagged for concerns and GRADE assessment was imprecise (OR = 0.68, 95% CI = (0.39 to 1.17)).

### New checks used by assessors

Assessors described eight checks which they used and which they felt were additional to the list of checks assessed in the study. Two of these – checking for trial registration, and checking the author list – were already covered by the primary list. Three others were variations of existing checks – checking the certification status of the ethical committee or institutional review board, looking at a related publication of a subgroup, and checking for consistency with the main article. Three were new: looking to see whether the authors exclusively worked together, checking whether the first author’s department had participated in other RCTs, and looking into the reported funder.

## Discussion

An extensive list of trustworthiness checks was assessed for their feasibility and impact by application to 95 RCTs in a sample of 50 Cochrane Reviews. The study allowed us to estimate how often each of the checks would be considered infeasible for routine use in systematic reviews, how often each would fail, and what the impact of applying the check would be on the estimates from meta-analysis. We found that, in the context of conducting a systematic review, the checks can be applied to identify problematic studies. Furthermore, the findings suggest that a substantial portion of meta-analyses would be left with no remaining RCTs if failed checks were used to identify and exclude problematic studies. Amongst those with remaining RCTs, there was a larger impact on precision than on the magnitude of effect estimates. The study also found that, following application of the checks, assessors frequently had concerns about the RCTs included in Cochrane Reviews, with “some concerns” being reported for 25% of studies, and ‘“serious concerns’” for a further 6%.

### Feasibility of the checks

A number of checks were deemed generally infeasible. For example, assessments which involved taking an author-wide view have been successfully implemented in particular cases (e.g. (8, 30, 31)), but were not considered feasible by assessors in the context of the current study. One possible reason is that these checks require additional data collection to find out more about the authors of a study, their research team, or their other publications. In a previous survey of experts, the need for a trustworthiness tool to be practical and not too burdensome was emphasised (15), and therefore checks which require the identification and comparison of additional studies are unlikely to be palatable. Other checks that were deemed infeasible include checking for evidence of copied work, including copied sample characteristics and results tables. Unless the copying is identified between RCTs that both happen to be included in the review, it is difficult to see how this sort of check would be practicable in the absence of automated solutions. Clearly, the results indicate that it would not be feasible to apply such a long list of checks routinely, as this took more than 3 hours for 42% of the trials.

### Identification of problematic studies

Failed checks are potential indicators of a problematic trial. Some checks failed for most RCTs. For example, the study agreed with previous work suggesting that many (in the present study, most) RCTs are not prospectively registered (32–34), and few make the underlying data available (35). In relation to registration, assessors were much more likely to have concerns about authenticity for studies that were not prospectively registered compared to those that were. This could indicate that lack of registration was influential in reaching an overall judgement, or rather that studies with other problematic features were less likely to be prospectively registered. Prospective registration is routinely considered in relation to reporting bias, and an important question to be resolved in the INSPECT-SR development process is whether there is additional value in considering prospective registration in the assessment of trustworthiness.

Our findings also indicate that some checks may be prone to misinterpretation or misapplication, which was suggested by high failure rates. In particular, several statistical checks proved challenging. For example, 20% of RCTs were considered to have ‘failed’ or ‘possibly failed’ a check looking to see whether results of statistical tests of outcomes were correct. Some of these failures might be attributable to the rounding of continuous variables in published articles; p-values obtained from rounded summary statistics can differ from those obtained from analysis of the underlying data, meaning the question of assessing consistency cannot just be assessed reproducing the test and looking for an exact match (36). Another example was checking differences in baseline variance between groups, which ‘“failed’” or ‘“possibly failed’” for 28% of RCTs. Assessors were directed to use an F test here. However, this test has an inflated type 1 error rate for skewed variables (37), such that rejection of the test assumptions may have been frequently mistaken for rejection of the hypothesis of equality of variances. Instances such as these may have led to unwarranted skepticism about a study’s authenticity in some instances. Although we did not detect associations between failure of these checks and concerns in post-hoc analyses, it remains possible that errors of this nature did sometimes occur, and may have influenced the overall assessment of a trial’s authenticity.

### Impact of identifying problematic studies on systematic reviews

The impact of removing RCTs flagged by these checks from meta-analyses may appear alarming; for example, removing RCTs for which assessors expressed concerns would result in 11 of 50 meta-analyses with no RCTs remaining. However, caution is needed here due to limitations introduced by our study design. We only assessed meta-analyses containing five or fewer trials in this study. Consequently, more than half contained only a single RCT, which is fewer than is typical (38). This will exaggerate the number of reviews with no remaining studies following trustworthiness assessment. Moreover, several other metrics, such as the change in point estimate and associated uncertainty, could only be evaluated in meta-analyses with at least one trial remaining following assessment. Due to the fact that many meta-analyses only included one trial initially, this subset will omit many of the meta-analyses with any trials flagged at all, causing the impact of the checks on these metrics to be understated.

In line with our expectations, there did not appear to be a clear association between Risk of Bias domains and overall trustworthiness assessment, reinforcing the premise that these frameworks are evaluating different aspects of trials. Many problematic studies appear to frequently describe perfectly sound methods (8). We were unable to ascertain whether there is any link between GRADE and trustworthiness assessment, as our estimate of the relationship was too imprecise. We suggest that trustworthiness assessment should be performed prior to Risk of Bias and GRADE assessments, because the value of assessing the internal or external validity of a problematic study is doubtful.

### Implications for development of INSPECT-SR and future directions

These observations have informed the development of the INSPECT-SR tool and accompanying guidance. The findings highlight the need for careful curation of the checks included in INSPECT-SR, and suggest that any statistical checks included in the tool would have to be accompanied by detailed guidance to enable their application, as well as to prevent misuse and misinterpretation. As technological solutions become available to facilitate some useful but difficult checks, they can become part of the tool implementation. As the role of automation, including artificial intelligence, is likely to expand in evidence synthesis, it will be important to examine how it might enable or hinder detection of problematic RCTs (39). For example, some checks, such as statistical checks, may be more amenable to automation than checks that require more content knowledge, such as the plausibility of participant recruitment or effect sizes.

Additional future directions informed by this study will be development of training for INSPECT-SR and tools that can be applied to individual patient data or observational study designs. Creating a searchable, open archive of trials that have been evaluated with INSPECT-SR will aid all systematic reviewers and users of trials. Lastly, although INSPECT-SR is being developed for use by systematic reviewers, adaptations of the tool could also be useful to journal editors or publishers who screen trials for research integrity problems.

## Conclusion

The study appears to reinforce the need for routine trustworthiness assessment in RCTs, suggesting that problematic studies in systematic reviews may not be infrequent, and are not detected by Risk of Bias assessment. Only two of the studies judged to be concerning had associated retraction or expression of concern notices at the point of assessment, highlighting the need to evaluate other features in order to identify these untrustworthy trials. The time taken to complete the full barrage of checks for each RCT was long, and would likely not be practicable in the context of a typical systematic review. The goal of subsequent stages of the INSPECT-SR project will be to identify a subset of these checks that are both feasible and useful, and to implement these in the form of a tool that can be implemented by systematic reviewers. The results from this study will be used to select checks for this purpose, alongside a Delphi study of experts and potential users of the tool. Both sets of results will be presented to experts at a series of consensus meetings, which will be used to determine the content of a draft version of INSPECT-SR. The draft version of the tool will then be tested in the assessment of RCTs, and feedback will be used to finalise the tool in early 2025.

## Declarations

JW, CH, GAA, LB, JJK declare funding from NIHR (NIHR203568) in relation to the current project. JW additionally declares Stats or Methodological Editor roles for BJOG, Fertility and Sterility, Reproduction and Fertility, Journal of Hypertension, and for Cochrane Gynaecology and Fertility. CH declares a Statistical Editor role for Cochrane Colorectal. GAA additionally declares a Statistical Reviewer role for the European Journal of Vascular and Endovascular Surgery. LB additionally declares a role as Academic Meta-Research Editor for PLoS Biology, and that The University of Colorado receives remuneration for service as Senior Research Integrity Editor, Cochrane. JJK additionally declares a Statistical Editor role for The BMJ. EF is employed by the Cochrane Collaboration and on the Editorial Board of Cochrane Evidence Synthesis and Methods. SL is an editor for Cochrane Gynaecology and Fertility, Human Reproduction, and Fertility and Sterility. TJL is the Deputy Editor in Chief of The Cochrane Library and is an employee of The Cochrane Collaboration. DNB is an associate editor for Research Quarterly for Exercise and Sport and a section editor for Communications in Kinesiology. NEO is a member of the Cochrane Editorial Board and holds an ERA-NET Neuron Co-Fund grant for a separate project. RR declares acting as an author and editor on Cochrane reviews. KS is an editor for Cochrane Gynaecology and Fertility, and Fertility and Sterility. MvW declares to be co-ordinating editor for Cochrane Gynaecology and Fertility and Cochrane Sexually Transmitted Infections, methodological editor for Human Reproduction Update and Editorial Editor for Fertility & Sterility. HT is Deputy Editor of The Lancet Gastroenterology & Hepatology and is an employee of Elsevier. SL received funding from the French National Research Agency (ANR-23-CE36-0006-01). AK is an editorial board member for BJGP Open. TLi serves as the Principal Investigator on a grant from the National Eye Institute, National Institutes of Health that funds the work of Cochrane Eyes and Vision US Project. She also acts as a sign-off editor for The Cochrane Library. ZM is supported by an NHMRC Investigator Grant 1195676. ZM is an associate Editor for BMC Medical Research Methodology and is on the Editorial Board for Clinical and Public Health Guidelines. RC is Editor-in-Chief at Meta-Psychology. CL is a work-package leader for the doctoral network MSCA-DN SHARE-CTD (HORIZON-MSCA-2022-DN-01 101120360), funded by the EU. CV received funding as part of the OSIRIS project (Open Science to Increase Reproducibility in Science); the OSIRIS (Open Science to Increase Reproducibility in Science) project has received funding from the EU (grant agreement No. 101094725). FN received funding from the French National Research Agency (ANR-23-CE36-0006-01), the French ministry of health and the French ministry of research. He is a work-package leader in the OSIRIS project (Open Science to Increase Reproducibility in Science). The OSIRIS project has received funding from the European Union’s Horizon Europe research and innovation programme under grant agreement No. 101094725. He is a work-package leader for the doctoral network MSCA-DN SHARE-CTD (HORIZON-MSCA-2022-DN-01 101120360), funded by the EU. DN declares having led/co-authored/co-authoring Cochrane Reviews. He also declares having been part of the Cochrane Convenes initiative organised by Cochrane to consider the issue of misinformation, its impact on the health evidence ecosystem and solutions to address it. LJ is the creator of the scrutiny package in R. WL is supported by an NHMRC Investigator grant (GNT2016729). RW is supported by an NHMRC Investigator Grant (2009767) and acts as a Deputy Editor for Human Reproduction, and an editorial board member for BJOG and Cochrane Gynaecology and Fertility. EF, SGTLa and RR declare employment by Cochrane. TLa additionally declares authorship of a chapter in the Cochrane Handbook for Systematic Reviews of Interventions and that he is a developer of standards for Cochrane intervention reviews (MECIR). AL is on the editorial board of BMC Medical Ethics.

## Ethical approval

The University of Manchester ethics decision tool was used on 30/09/22. Ethical approval was not required for this study, since it involved appraisal of published research.

## Data Availability

The study dataset and R code is available at https://osf.io/9pyw2/.

