## Supplementary tables for "Assessing the feasibility and impact of clinical trial trustworthiness checks via an application to Cochrane Reviews: Stage 2 of the INSPECT-SR project"

**Supplementary Material**

**Section A: List of checks and explanations**

**Section B: Additional characteristics of the sample**

**Section C: Check responses vs overall concerns**

**Section D: Impact of removing RCTs flagged by each check from meta-analyses**

**Section A. List of checks and explanations**

**Inspecting the results in the paper (28 checks)**

| **Number** | **Item** | **Explanation** |
| --- | --- | --- |
| 1 | Are any baseline data implausible with respect to magnitude, frequency, or variance? | Look at the summary statistics relating to the baseline characteristics. This may include measures of central tendency (e.g. means, medians), measures of dispersion (e.g. standard deviation, interquartile range, range), and frequencies/ proportions. Consider whether the values are plausible. For example, are measures of dispersion unusually small? For standard deviations, consider the possibility that authors may have unintentionally reported standard errors. |
| 2 | Is the number of participant withdrawals compatible with the disease, age and timeline? | Some empirical work has suggested patterns of withdrawals tend to be unusual in inauthentic trials. Possible indicators of untrustworthiness include low numbers of withdrawals and very similar or equal numbers of withdrawals across study arms. Consider the plausibility of the number of withdrawals given the context. |
| 3 | Are subgroup means incompatible with those for the whole cohort? | If means for an exhaustive and exclusive set of subgroups are reported, the mean for the whole group can be calculated. Consequently, the reported group mean can be checked against the value calculated from the subgroup means. An Excel sheet ‘Subgroup means’ can be used to perform this calculation, in the Statistics Checks file. |
| 4 | Are the reported summary data compatible with the reported range? | If a range is reported, consider whether any other reported values contradict this (e.g. by falling outside of the range) |
| 5 | Are correct units reported? | Check to see if any results have incorrect units, which could be an indication that results for a different variable were copied but authors forgot to amend the units. |
| 6 | Are calculations of proportions and percentages correct? | Check that the calculated proportions can be reproduced from reported frequencies and group sizes. Suggest performing this for at least several incidental results (fabricators may pay less attention to checking incidental compared to important results in the paper). |
| 7 | Are numbers of participants correct and consistent throughout the publication? | Check that numbers of participants are consistent throughout, including in tables and figures, or that any discrepancies can be accounted for (for example, due to the exclusion of patient withdrawals from analysis) |
| 8 | Are there any discrepancies between data reported in figures, tables and text? | Check that results shown are consistent between text, tables, figures. |
| 9 | Are any outcome data, including estimated treatment effects, implausible? | Consider whether the reported outcomes, including reported treatment effects, are plausible. It is important to remember that estimated treatment effects are merely *estimates*, and that it is not unusual to obtain large estimates when conditions are such that the treatment effect cannot be estimated precisely. The estimate should not be considered in isolation from any corresponding statistical inference in the article, such as p-values or 95% confidence intervals. A large estimate of treatment effect might be considered worthy of further inspection if it is accompanied by a narrow 95% CI or very small p-value. |
| 10 | Are baseline statistical tests correct? | Some statistical tests may be reproduced from summaries reported in the paper. For example, t-tests may be reproduced using reported means, standard deviations, and group sizes, and chi-squared tests may be reproduced from reported frequencies and group sizes. These can be checked against results reported in the paper. The Excel workbook ‘Table 1 extraction’ in the Statistics Checks file can be used to reproduce t-tests (under the assumption of equal variances) and chi-squared tests. |
| 11 | Are statistical tests of outcomes correct? | Some statistical tests may be reproduced from summaries reported in the paper. For example, t-tests may be reproduced using reported means, standard deviations, and group sizes, and chi-squared tests may be reproduced from reported frequencies and group sizes. These can be checked against results reported in the paper. These can be checked using the ‘Outcome extraction’ sheet of the Statistics Checks file. |
| 12 | Are differences in variances in baseline variables between randomised groups plausible? | Variances in baseline characteristics should not differ by more than would be expected by chance under random allocation. This can be assessed using an F-test, for example. The Excel workbook ‘Table 1 extraction’ in the Statistics Checks file can be used to perform an F-test of the equality of variances. |
| Stouffer p-value |  | It has been proposed that summaries of baseline data from RCTs may be used to detect problematic studies. One possibility is that baseline characteristics might be more similar or more different than would be expected under randomisation. Excessive similarity or difference could be judged qualitatively, or quantitatively. Methods for quantitative assessment include the Stouffer method, which can be used to calculate a combined p-value from the baseline table. Combined p-values below 5% or above 95% have been used to indicate excessive difference or similarity. Threats to the validity of this approach include: methods of randomisation designed to improve balance in prognostic variables, such as minimisation and stratified randomisation; rounding of summary statistics; the non-independence of tests of variables in a given study; non-normality of continuous variables. The method also does not apply to categorical baseline variables.  If baseline data and any p-values reported by the authors are entered into the Excel spreadsheet ‘Table 1 extraction’ in the Statistics Checks file then the Stouffer combined p-value will be calculated on the basis of the continuous variables only. Where p-values are reported by the authors, these will be used for the calculation. Where no p-values are reported by the authors, the spreadsheet will use the p-value obtained from a t-test. |
| 13 | Are any of the baseline data excessively similar between randomized groups? | We recommend first making a judgement as to whether the baseline characteristics appear excessively similar, and supplementing this using the Stouffer p-value (e.g. a Stouffer p-value >= 0.95 has been used to suggest excessive similarity). Remember that some treatment allocation methods are designed to improve balance on baseline variables, and in this context similarity should not be considered suspicious. |
| 14 | Are any of the baseline data excessively different between randomised groups? | We recommend first making a judgement as to whether the baseline characteristics appear excessively different. This can be supplemented by the Stouffer p-value (a p-value <= 0.05 has been used to suggest excessive difference). The Stouffer p-value is based only on continuous variables, and imbalances in individual variables could also be considered (for example, using p-values reported by the authors or values obtained from the ‘Table 1 extraction’ sheet in the Statistics Checks file. |
| 15 | Are the summary outcome data identical or nearly identical across study groups? | Check whether outcome data are the same or very similar in the study groups. |
| 16 | Are there any discrepancies between the values for percentage and absolute change? | If both absolute and percentage changes are reported for the same variable, these can be compared to check that they match. |
| 17 | Are there any discrepancies between reported data and participant inclusion criteria? | Participant characteristics can be compared to study inclusion criteria for consistency. |
| 18 | Are the variances in biological variables surprisingly consistent over time? | Where one variable is measured on multiple occasions, check to see whether measures of variance (e.g. standard deviation) remain similar at each timepoint. Similarity could indicate inauthenticity (and so the check would be failed). |
| 19 | Are results internally consistent? | Some combinations of results are not possible. For example, it is not possible to have more birth events (the birth of at least one child) than pregnancies. Check that no contradictions are implied by the results. |
| 20 | Are coefficients of variation unusually similar when calculated across variables reported in the paper? | A coefficient of variation can be calculated as 100*SD/mean. It has been proposed that for some inauthentic studies, coefficients of variation calculated for different variables in the study might be similar. This is calculated by the Excel sheet ‘Table 1 extraction’ in the Statistics Checks file. |
| 21 | Is the amount of missing data plausible? | Is the amount of missing data in the study plausible given the context? Is it lower than would be considered plausible? |
| 22 | Are the results substantially divergent from the results of multiple other studies in meta-analysis? | It has been proposed that, if a study has highly discordant results compared to other similar studies which we are considering for pooling in a meta-analysis, then this might be a sign that the study could be untrustworthy. Individual studies may produce imprecise estimates of treatment effect, and so a particularly large or small estimate would not be sufficient to suggest problems; there may nonetheless be considerable overlap between the confidence intervals from that study and others. If the confidence interval does not overlap with others in the meta-analysis, this may highlight the result as being discrepant. We recommend looking at the forest plot corresponding to the meta-analysis in the Cochrane Review, to consider whether the result is discrepant. |
| 23 | Are non-first digits compatible with a genuine measurement process? | It has been suggested that statistical examination  of numbers that are of inconsequential meaning can highlight inauthentic data. Inconsequential digits can be compared to a uniform distribution. For example, when values are recorded to several decimal places, the values in the final decimal place position may be extracted and compared to a uniform distribution, for example using a chi-squared test. The Excel sheet ‘Mossiman last digit chisquare’ in the Statistics Checks file extracts the last reported decimal place value and performs a chi-squared test. This check would be failed if the test is significant. |
| 24 | Are the variances of integer data possible? | The mathematical (im)possibility of a reported variance for discrete data (e.g. ordinal variables) can be considered. GRIMMER can be used for this purpose. A web-based implementation of this check is available at <http://www.prepubmed.org/grimmer/> |
| 25 | Are the means of integer data possible? | The mathematical possibility of a reported mean for discrete data can be considered. This includes ordinal data, but also summaries of binary data when reported as percentages or proportions. GRIM can be used for this purpose. The Excel sheet ‘GRIM test’ in the Statistics Checks file can be used for this purpose. R users can implement this check on multiple variables at once using the grim_map_seq() function of the scrutiny package: <https://cran.r-project.org/web/packages/scrutiny/scrutiny.pdf> |
| 26 | Is there heterogeneity across studies in degree of imbalance in baseline characteristics (in meta-analysis) – only once per review | This check is performed across all studies in a meta-analysis, rather than for a single trial. Summaries of one or more key baseline prognostic characteristics (e.g. age) are extracted for each study group in each study. The proposal is to consider heterogeneity in the degree of balance across studies, for example using a Q statistic, or I-squared. Differences in randomisation method between studies need to be taken into account. For example, we would expect baseline variables to be more similar, in trials which used a method of allocation intended to increase balance of baseline covariates (such as stratified randomisaiton or minimisation). This check can be implemented using the ‘Meta-analysis heterogeneity’ sheet in the Statistics Checks file. The result of this check should be the same for all of the trials you are assessing in a meta-analysis. |
| 27 | Are integer data simulated from reported summary statistics plausible? | Datasets may be simulated from reported summary statistics for discrete variables, and visualised as an aid in considering the plausibility of the data. This may be performed using SPRITE: [rSPRITE beta 0.18 (shinyapps.io)](https://steamtraen.shinyapps.io/rsprite/) |
| 28 | Are important features missing from the paper? | Check to see whether expected features are missing, such as figures. |

S Table 1: Checks in the domain ‘Inspecting results in the paper’

**Inspecting the research team and their work (19 checks)**

| **Number** | **Item** | **Explanation** |
| --- | --- | --- |
| 29 | Check whether withdrawal and loss to follow-up in multiple trials by the same author are consistent with the expected (random) binomial distribution | Multiple fabricated trials from the same author(s) might display similar unusual features, such as consistently low dropout or withdrawal rates, or very similar dropout/ withdrawal rates between arms. This check would require the identification of multiple studies from the same author however, and so may not be feasible. |
| 30 | Are contributorship statements present? | Self-explanatory, although rationale not entirely clear. If not present, rate this as ‘failed’. |
| 31 | Are contributorship statements complete? | Self-explanatory, although rationale not entirely clear. If not complete, rate this as ‘failed’. |
| 32 | Have the data been published elsewhere by the research team in an illegitimate fashion? | Have the results been published elsewhere? Does it appear that results have been published in duplicate in an illegitimate fashion (this would not be the case for conference abstracts or preprints, for example)? |
| 33 | Are duplicate-reported data consistent between publications? | Where a study is reported in several places (e.g. conference abstract, preprint, journal article) are results consistent between publications? This would require identifying additional reports, so may not be feasible. |
| 34 | Are relevant methods consistent between publications? | Where a study is reported in several places (e.g. conference abstract, preprint, journal article) are methods consistent between publications? This would require identifying additional reports, so may not be feasible. |
| 35 | Is any duplicate reporting acknowledged or explained? | If results have been published in duplicate (not including conference abstracts or preprints, for example) is this acknowledged or explained in either publication? This would require identifying additional reports, so may not be feasible. |
| 36 | Is there evidence of duplication of figures? | Is there evidence that any figures in the article have been copied from elsewhere? This may not be feasible. |
| 37 | Does the statistics methods section use generic language, suggesting lack of expert statistical input? | Generic text may indicate lack of collaboration with a statistical expert, which could be an incidental feature of problematic studies. |
| 38 | Is the distribution of non-first digits in manuscripts from one author compatible with a genuine measurement process? | It has been suggested that statistical examination  of numbers that are of inconsequential meaning can highlight inauthentic data. Inconsequential digits can be compared to a uniform distribution. For example, when values are recorded to several decimal places, the values in the final decimal place position may be extracted and compared to a uniform distribution, for example using a chi-squared test.  The proposal here is to perform this assessment across multiple studies from the same author. This would require identification of multiple studies from one author, so may not be feasible. |
| 39 | Does consideration of other studies from members of the research team highlight causes for concern (including expressions of concern, relevant post-publication amendment, or critical retraction)? | A search of author names can be performed using Retraction Watch (<http://retractiondatabase.org/RetractionSearch.aspx>?) and PubPeer (<https://pubpeer.com/> ) to identify concerns with published work. We recommend performing this search for at least the first and last authors. |
| 40 | Is the standard deviation of summary statistics in multiple studies by same authors plausible (when compared to simulated or bootstrapped data?) | It has been suggested that summary statistics from fabricated studies from the same author might tend to be more similar than would be expected in authentic data. The proposal is to compare the variation in summary statistics from multiple papers to bootstrapped or simulated summary statistics. This may not be feasible, as it would require that multiple studies from one author are identified, and would require advanced statistical expertise to perform the analysis. |
| 41 | Do all authors meet criteria for authorship? | This might be difficult to assess. There may be no other way to consider this than to consider the contributorship statement. |
| 42 | Is authorship of related papers consistent? | Where multiple reports relating to the same study exist, we can check whether authorship is consistent across publications. |
| 43 | Are the authors on staff of institutions they list? | False affiliations may be used. Searching for the institutional profile of authors can be performed to verify their affiliation (or that they have another relevant affiliation, allowing for the possibility they may have moved institutions). Recommend performing this for at least the first and last author. |
| 44 | Do any authors have a professorial title but no other publications on PubMed? | False titles might be used. It would be unusual for a senior researcher to have no research record. The name(s) can be searched on PubMed. This could be performed for senior authors in key authorship positions (e.g. last author). |
| 45 | Can co-authors attest to the reliability of the paper? | If co-authors cannot confirm the veracity of the data in the study, this may be a cause for concern. Contacting co-authors is not expected in the context of the current study, so this is not to be assessed. |
| 46 | Given the nature of the study, does the author list make sense? - e.g.does a simple study have dozens of authors from different institutions and with diverse expertise? | Consider whether the author list is plausible (e.g. a large, complex study may have too few authors, or vice versa). |
| 47 | In which country was the study conducted? | Record the country/ countries in which the study took place. It may be that research conducted in circumstances where this is less research governance is more susceptible to problems. The current study might offer some preliminary insight into this. |

S Table 2: Checks in the domain ‘Inspecting the research team and their work’

**Inspecting conduct, governance, and transparency (22 checks)**

| **Number** | **Item** | **Explanation** |
| --- | --- | --- |
| 48 | Is the grant funding number identical to the number in unrelated studies? | A false grant funding number may be provided. This can be searched to see if it corresponds to an unrelated study. |
| 49 | Is a funding source reported? | Record whether a funding source is reported in the manuscript. |
| 50 | Is the volume of work reported by research group plausible, including that indicated by concurrent studies from the same group? | This would require an assessment of the volume of total work conducted by the group, including other studies. It may not be feasible to assess this in the context of the present study. |
| 51 | Is the reported staffing adequate for the study conduct as reported? | Consider whether the reported study would be feasible given the reported staffing. |
| 52 | Is the recruitment of participants plausible within the stated time frame for the research? | Consider whether the recruitment is plausible in the study duration. |
| 53 | Is the recruitment of participants plausible considering the epidemiology of the disease in the area of the study location? | Consider whether the overall number of participants recruited is plausible given the prevalence of the study condition. |
| 54 | Is the interval between study completion and manuscript submission plausible? | Consider the plausibility of the study end date in relation to the manuscript submission date. |
| 55 | Is there evidence that the work has been approved by a specific, recognized committee? (ethics) | Search for evidence of ethical approval, including a search for the named committee. |
| 56 | Are there any concerns about unethical practice? | Consider whether any aspect of the study could be considered unethical. |
| 57 | Could the study plausibly be completed as described? | Consider the plausibility of completing the study. |
| 58 | Are the study methods plausible, at the location specified? | Consider the plausibility of the methods employed in the study, at the scale described. |
| 59 | Are the locations where the research took place specified, and is this information plausible? | Consider the plausibility that the study took place at the location described. |
| 60 | Do the authors agree to share individual participant data? | We will not assess this in the present study. |
| 61 | Are the data publically available? | This check would be failed if the data do not appear to be publically available. |
| 62 | Are additional patient data recorded in patient case records beyond what is reported in the paper? | This is unlikely to be feasible in the context of the present study. |
| 63 | Does the trial registration number refer to other studies? | A fake trial registration number may be used. Search for the registration number to see whether it refers ot another study. |
| 64 | Has the study been prospectively registered? | Check whether the study has been prospectively registered. This will usually involve searching on the trial registration number and comparing the registration date to the study start date. |
| 65 | Are details such as dates and study methods in the publication consistent with those in the registration documents? | Check consistency of dates on the registration and published paper. |
| 66 | Do authors cooperate with requests for information? | This will not be assessed in the present study. |
| 67 | Do authors provide satisfactory responses to requests? | This will not be assessed in the present study |
| 68 | Was the time between submission to acceptance reasonable? | Consider whether the time between submission and acceptance of the manuscript was too short, suggesting that the paper may not have undergone a thorough peer review and editorial assessment. |
| 69 | Is the procedure of the study aligned with local legislations? | This may be difficult to assess without knowledge of the local context. |

S Table 3: Checks in the domain ‘Inspecting conduct, governance and transparency’

**Inspecting text and publication details (7 checks)**

| **Number** | **Item** | **Explanation** |
| --- | --- | --- |
| 70 | Are there typographical errors? | Check whether there are typos in the work. |
| 71 | Has the study been retracted or does it have an expression of concern, a relevant post-publication amendment, a critical Retraction Watch or PubPeer comment or has been previously excluded from a systematic review? | Check to see whether this study has an expression of concern or retraction notice on the journal website, or relevant critical comments on Retraction Watch (<http://retractiondatabase.org/RetractionSearch.aspx>? ) or PubPeer (<https://pubpeer.com/> ). |
| 72 | Is there evidence of copied work, such as duplicated or partially duplicated tables? | Fabricators might copy tables from other articles, in part or in whole. This would require identification of the original tables however, and may not be feasible in the context of the present study. |
| 73 | Is there evidence of text reuse (cutting and pasting text between papers), including text that is inconsistent with the study? | Text may have been copied from other studies, and this may include unintentional copying of phrases which are not consistent with the study. Check whether any such phrases are apparent. |
| 74 | Is there evidence of automatically-generated text? | Consider whether any phrases are unusual or unnatural, which may be indicative of automatically-generated text. |
| 75 | Was the study published in journal from a list of predatory/ low quality journals? | There is no single agreed-upon definition of a predatory journal, but commonly-accepted characteristics include publication charges, and no substantive peer review or editorial process. There is no single, authoritative list of predatory journals, but we recommend making a judgement based on your knowledge of the field where possible. |
| 76 | Is there evidence of manipulation or duplication of images? | In some trials, there may be evidence of manipulated figures. This may include duplication and copying within the figure itself. Other signs of problems within a figure may include implausible repeating sequences, or clear discrepancies between the figure and reported results (such as discrepancies between the number of observations displayed on a scatterplot and reported number of participants). |

S Table 4: Checks in the domain ‘Inspecting text and publication details’

**Section B – Additional characteristics of the sample**

| Cochrane Acute Respiratory Infections Group | 2 (4%) |
| --- | --- |
| Cochrane Airways Group | 1 (2%) |
| Cochrane Back and Neck Group | 2 (4%) |
| Cochrane Breast Cancer Group | 2 (4%) |
| Cochrane Common Mental Disorders Group | 1 (2%) |
| Cochrane Consumers and Communication Group | 2 (4%) |
| Cochrane Cystic Fibrosis and Genetic Disorders Group | 2 (4%) |
| Cochrane Developmental, Psychosocial and Learning Problems Group | 4 (8%) |
| Cochrane Drugs and Alcohol Group | 2 (4%) |
| Cochrane ENT Group | 2 (4%) |
| Cochrane Epilepsy Group | 1 (2%) |
| Cochrane Eyes and Vision Group | 3 (6%) |
| Cochrane Gut Group | 2 (4%) |
| Cochrane Gynaecology and Fertility Group | 2 (4%) |
| Cochrane Haematology Group | 1 (2%) |
| Cochrane Hypertension Group | 1 (2%) |
| Cochrane Incontinence Group | 2 (4%) |
| Cochrane Musculoskeletal Group | 5 (10%) |
| Cochrane Neonatal Group | 3 (6%) |
| Cochrane Pregnancy and Childbirth Group | 4 (8%) |
| Cochrane Public Health Group | 1 (2%) |
| Cochrane Skin Group | 1 (2%) |
| Cochrane Work Group | 1 (2%) |
| Cochrane Wounds Group | 3 (6%) |

S Table 5: Cochrane Groups for the 50 assessed Cochrane Reviews. Frequency (%)

| Number of participants in assessed RCT: median (IQR) | 71 (40 to 174) |
| --- | --- |
| Country in which RCT was conducted: n (%) |  |
| Multiple countries | 15 (16) |
| USA | 15 (16) |
| UK | 9 (9) |
| Australia | 7 (7) |
| India | 7 (7) |
| Iran | 7 (7) |
| Italy | 7 (7) |
| Japan | 4 (4) |
| Brazil | 3 (3) |
| Spain | 3 (3) |
| France | 2 (2) |
| Germany | 2 (2) |
| Israel | 2 (2) |
| Netherlands | 2 (2) |
| Norway | 2 (2) |
| Saudi Arabia | 2 (2) |
| China | 1 (1) |
| Denmark | 1 (1) |
| Ireland | 1 (1) |
| Pakistan | 1 (1) |
| South Africa | 1 (1) |
| South Korea | 1 (1) |

S Table 6: Characteristics of 95 RCTs assessed in the study

**Section B – Check responses**

| Check | n | Not Feasible | Passed | Possible Fail | Fail | Not Feasible (%) | Passed (%) | Possible Fail (%) | Fail (%) |
| --- | --- | --- | --- | --- | --- | --- | --- | --- | --- |
| check1 | 95 | 18 | 67 | 6 | 4 | 19 | 71 | 6 | 4 |
| check2 | 95 | 8 | 74 | 10 | 3 | 8 | 78 | 11 | 3 |
| check3 | 94 | 76 | 14 | 3 | 1 | 81 | 15 | 3 | 1 |
| check4 | 93 | 35 | 53 | 2 | 3 | 38 | 57 | 2 | 3 |
| check5 | 94 | 12 | 78 | 3 | 1 | 13 | 83 | 3 | 1 |
| check6 | 94 | 19 | 64 | 7 | 4 | 20 | 68 | 7 | 4 |
| check7 | 95 | 2 | 85 | 4 | 4 | 2 | 89 | 4 | 4 |
| check8 | 95 | 0 | 86 | 3 | 6 | 0 | 91 | 3 | 6 |
| check9 | 95 | 8 | 76 | 10 | 1 | 8 | 80 | 11 | 1 |
| check10 | 93 | 49 | 30 | 7 | 7 | 53 | 32 | 8 | 8 |
| check11 | 94 | 28 | 46 | 14 | 6 | 30 | 49 | 15 | 6 |
| check12 | 93 | 24 | 43 | 18 | 8 | 26 | 46 | 19 | 9 |
| check13 | 94 | 22 | 62 | 7 | 3 | 23 | 66 | 7 | 3 |
| check14 | 93 | 19 | 62 | 9 | 3 | 20 | 67 | 10 | 3 |
| check15 | 95 | 9 | 80 | 6 | 0 | 9 | 84 | 6 | 0 |
| check16 | 93 | 67 | 23 | 1 | 2 | 72 | 25 | 1 | 2 |
| check17 | 95 | 12 | 81 | 2 | 0 | 13 | 85 | 2 | 0 |
| check18 | 95 | 56 | 37 | 2 | 0 | 59 | 39 | 2 | 0 |
| check19 | 93 | 18 | 69 | 4 | 2 | 19 | 74 | 4 | 2 |
| check20 | 93 | 29 | 55 | 9 | 0 | 31 | 59 | 10 | 0 |
| check21 | 95 | 7 | 71 | 14 | 3 | 7 | 75 | 15 | 3 |
| check22 | 94 | 22 | 61 | 4 | 7 | 23 | 65 | 4 | 7 |
| check23 | 94 | 22 | 60 | 5 | 7 | 23 | 64 | 5 | 7 |
| check24 | 91 | 58 | 30 | 0 | 3 | 64 | 33 | 0 | 3 |
| check25 | 93 | 45 | 39 | 3 | 6 | 48 | 42 | 3 | 6 |
| check26 | 85 | 55 | 28 | 2 | 0 | 65 | 33 | 2 | 0 |
| check27 | 93 | 52 | 35 | 4 | 2 | 56 | 38 | 4 | 2 |
| check28 | 95 | 2 | 64 | 16 | 13 | 2 | 67 | 17 | 14 |
| check29 | 94 | 79 | 14 | 1 | 0 | 84 | 15 | 1 | 0 |
| check30 | 95 | 0 | 29 | 4 | 62 | 0 | 31 | 4 | 65 |
| check31 | 95 | 17 | 24 | 4 | 50 | 18 | 25 | 4 | 53 |
| check32 | 95 | 63 | 30 | 0 | 2 | 66 | 32 | 0 | 2 |
| check33 | 93 | 71 | 17 | 2 | 3 | 76 | 18 | 2 | 3 |
| check34 | 93 | 73 | 17 | 3 | 0 | 78 | 18 | 3 | 0 |
| check35 | 93 | 83 | 8 | 0 | 2 | 89 | 9 | 0 | 2 |
| check36 | 93 | 77 | 13 | 2 | 1 | 83 | 14 | 2 | 1 |
| check37 | 95 | 4 | 61 | 19 | 11 | 4 | 64 | 20 | 12 |
| check38 | 92 | 83 | 8 | 1 | 0 | 90 | 9 | 1 | 0 |
| check39 | 95 | 13 | 72 | 5 | 5 | 14 | 76 | 5 | 5 |
| check40 | 95 | 94 | 1 | 0 | 0 | 99 | 1 | 0 | 0 |
| check41 | 94 | 59 | 26 | 3 | 6 | 63 | 28 | 3 | 6 |
| check42 | 90 | 66 | 22 | 2 | 0 | 73 | 24 | 2 | 0 |
| check43 | 94 | 9 | 79 | 4 | 2 | 10 | 84 | 4 | 2 |
| check44 | 94 | 17 | 70 | 4 | 3 | 18 | 74 | 4 | 3 |
| check45 | NA | NA | NA | NA | NA | NA | NA | NA | NA |
| check46 | 95 | 0 | 79 | 14 | 2 | 0 | 83 | 15 | 2 |
| check47 | NA | NA | NA | NA | NA | NA | NA | NA | NA |
| check48 | 95 | 63 | 25 | 4 | 3 | 66 | 26 | 4 | 3 |
| check49 | 94 | 0 | 56 | 0 | 38 | 0 | 60 | 0 | 40 |
| check50 | 95 | 37 | 51 | 7 | 0 | 39 | 54 | 7 | 0 |
| check51 | 95 | 24 | 62 | 9 | 0 | 25 | 65 | 9 | 0 |
| check52 | 93 | 17 | 71 | 2 | 3 | 18 | 76 | 2 | 3 |
| check53 | 95 | 10 | 80 | 5 | 0 | 11 | 84 | 5 | 0 |
| check54 | 95 | 29 | 56 | 8 | 2 | 31 | 59 | 8 | 2 |
| check55 | 95 | 4 | 67 | 20 | 4 | 4 | 71 | 21 | 4 |
| check56 | 94 | 12 | 71 | 11 | 0 | 13 | 76 | 12 | 0 |
| check57 | 95 | 5 | 85 | 3 | 2 | 5 | 89 | 3 | 2 |
| check58 | 95 | 2 | 92 | 1 | 0 | 2 | 97 | 1 | 0 |
| check59 | 95 | 2 | 77 | 11 | 5 | 2 | 81 | 12 | 5 |
| check60 | NA | NA | NA | NA | NA | NA | NA | NA | NA |
| check61 | 94 | 9 | 9 | 10 | 66 | 10 | 10 | 11 | 70 |
| check62 | 92 | 90 | 2 | 0 | 0 | 98 | 2 | 0 | 0 |
| check63 | 95 | 55 | 14 | 0 | 26 | 58 | 15 | 0 | 27 |
| check64 | 95 | 12 | 31 | 9 | 43 | 13 | 33 | 9 | 45 |
| check65 | 93 | 48 | 35 | 5 | 5 | 52 | 38 | 5 | 5 |
| check66 | NA | NA | NA | NA | NA | NA | NA | NA | NA |
| check67 | NA | NA | NA | NA | NA | NA | NA | NA | NA |
| check68 | 93 | 40 | 38 | 12 | 3 | 43 | 41 | 13 | 3 |
| check69 | 94 | 69 | 24 | 0 | 1 | 73 | 26 | 0 | 1 |
| check70 | 95 | 0 | 88 | 4 | 3 | 0 | 93 | 4 | 3 |
| check71 | 95 | 6 | 87 | 1 | 1 | 6 | 92 | 1 | 1 |
| check72 | 95 | 58 | 37 | 0 | 0 | 61 | 39 | 0 | 0 |
| check73 | 94 | 26 | 64 | 2 | 2 | 28 | 68 | 2 | 2 |
| check74 | 95 | 15 | 80 | 0 | 0 | 16 | 84 | 0 | 0 |
| check75 | 95 | 6 | 84 | 3 | 2 | 6 | 88 | 3 | 2 |
| check76 | 94 | 15 | 76 | 3 | 0 | 16 | 81 | 3 | 0 |

S Table 7: Summary of assessments for each check

**Section C – Check responses vs overall concerns**

|  | Overall Concern | | | |  |
| --- | --- | --- | --- | --- | --- |
|  | Don't know | No | Some concerns | Serious concerns | p-value |
| Check1 |  |  |  |  | 1.12E-05 |
| Pass | 1 | 49 | 15 | 1 |  |
| % | 2 | 74 | 23 | 2 |  |
| Fail/Possible fail | 0 | 4 | 1 | 5 |  |
| % | 0 | 40 | 10 | 50 |  |
| Check2 |  |  |  |  | 0.004766 |
| Pass | 2 | 52 | 17 | 2 |  |
| % | 3 | 71 | 23 | 3 |  |
| Fail/Possible fail | 0 | 5 | 4 | 4 |  |
| % | 0 | 38 | 31 | 31 |  |
| Check3 |  |  |  |  | 0.944208 |
| Pass | 0 | 12 | 1 | 0 |  |
| % | 0 | 92 | 8 | 0 |  |
| Fail/Possible fail | 0 | 4 | 0 | 0 |  |
| % | 0 | 100 | 0 | 0 |  |
| Check4 |  |  |  |  | 0.914683 |
| Pass | 0 | 37 | 11 | 4 |  |
| % | 0 | 71 | 21 | 8 |  |
| Fail/Possible fail | 0 | 3 | 1 | 1 |  |
| % | 0 | 60 | 20 | 20 |  |
| Check5 |  |  |  |  | 0.137349 |
| Pass | 1 | 51 | 21 | 4 |  |
| % | 1 | 66 | 27 | 5 |  |
| Fail/Possible fail | 0 | 2 | 0 | 2 |  |
| % | 0 | 50 | 0 | 50 |  |
| Check6 |  |  |  |  | 0.992338 |
| Pass | 3 | 43 | 13 | 4 |  |
| % | 5 | 68 | 21 | 6 |  |
| Fail/Possible fail | 0 | 7 | 3 | 1 |  |
| % | 0 | 64 | 27 | 9 |  |
| Check7 |  |  |  |  | 0.263677 |
| Pass | 3 | 55 | 22 | 4 |  |
| % | 4 | 65 | 26 | 5 |  |
| Fail/Possible fail | 0 | 4 | 2 | 2 |  |
| % | 0 | 50 | 25 | 25 |  |
| Check8 |  |  |  |  | 6.10E-05 |
| Pass | 4 | 57 | 22 | 2 |  |
| % | 5 | 67 | 26 | 2 |  |
| Fail/Possible fail | 0 | 3 | 2 | 4 |  |
| % | 0 | 33 | 22 | 44 |  |
| Check9 |  |  |  |  | 1.92E-06 |
| Pass | 2 | 54 | 18 | 1 |  |
| % | 3 | 72 | 24 | 1 |  |
| Fail/Possible fail | 0 | 2 | 4 | 5 |  |
| % | 0 | 18 | 36 | 45 |  |
| Check10 |  |  |  |  | 0.176726 |
| Pass | 2 | 19 | 4 | 4 |  |
| % | 7 | 66 | 14 | 14 |  |
| Fail/Possible fail | 0 | 5 | 8 | 1 |  |
| % | 0 | 36 | 57 | 7 |  |
| Check11 |  |  |  |  | 0.634846 |
| Pass | 2 | 34 | 7 | 2 |  |
| % | 4 | 76 | 16 | 4 |  |
| Fail/Possible fail | 0 | 11 | 6 | 3 |  |
| % | 0 | 55 | 30 | 15 |  |
| Check12 |  |  |  |  | 0.467726 |
| Pass | 1 | 30 | 8 | 3 |  |
| % | 2 | 71 | 19 | 7 |  |
| Fail/Possible fail | 0 | 12 | 11 | 3 |  |
| % | 0 | 46 | 42 | 12 |  |
| Check13 |  |  |  |  | 0.924749 |
| Pass | 1 | 40 | 16 | 4 |  |
| % | 2 | 66 | 26 | 7 |  |
| Fail/Possible fail | 0 | 5 | 3 | 2 |  |
| % | 0 | 50 | 30 | 20 |  |
| Check14 |  |  |  |  | 0.449566 |
| Pass | 1 | 41 | 16 | 3 |  |
| % | 2 | 67 | 26 | 5 |  |
| Fail/Possible fail | 0 | 5 | 4 | 3 |  |
| % | 0 | 42 | 33 | 25 |  |
| Check15 |  |  |  |  | 0.114202 |
| Pass | 1 | 52 | 22 | 4 |  |
| % | 1 | 66 | 28 | 5 |  |
| Fail/Possible fail | 0 | 2 | 2 | 2 |  |
| % | 0 | 33 | 33 | 33 |  |
| Check16 |  |  |  |  | 0.951233 |
| Pass | 0 | 18 | 3 | 1 |  |
| % | 0 | 82 | 14 | 5 |  |
| Fail/Possible fail | 0 | 2 | 1 | 0 |  |
| % | 0 | 67 | 33 | 0 |  |
| Check17 |  |  |  |  | 0.172656 |
| Pass | 1 | 55 | 18 | 6 |  |
| % | 1 | 69 | 23 | 8 |  |
| Fail/Possible fail | 0 | 0 | 2 | 0 |  |
| % | 0 | 0 | 100 | 0 |  |
| Check18 |  |  |  |  | 0.717523 |
| Pass | 0 | 27 | 9 | 1 |  |
| % | 0 | 73 | 24 | 3 |  |
| Fail/Possible fail | 0 | 1 | 1 | 0 |  |
| % | 0 | 50 | 50 | 0 |  |
| Check19 |  |  |  |  | 8.09E-05 |
| Pass | 4 | 44 | 18 | 2 |  |
| % | 6 | 65 | 26 | 3 |  |
| Fail/Possible fail | 0 | 1 | 1 | 4 |  |
| % | 0 | 17 | 17 | 67 |  |
| Check20 |  |  |  |  | 0.878729 |
| Pass | 1 | 34 | 17 | 2 |  |
| % | 2 | 63 | 31 | 4 |  |
| Fail/Possible fail | 0 | 4 | 5 |  |  |
| % | 0 | 44 | 56 | 0 |  |
| Check21 |  |  |  |  | 0.014548 |
| Pass | 3 | 50 | 15 | 2 |  |
| % | 4 | 71 | 21 | 3 |  |
| Fail/Possible fail | 0 | 7 | 6 | 4 |  |
| % | 0 | 41 | 35 | 24 |  |
| Check22 |  |  |  |  | 0.03667 |
| Pass | 1 | 44 | 14 | 1 |  |
| % | 2 | 73 | 23 | 2 |  |
| Fail/Possible fail | 2 | 3 | 4 | 2 |  |
| % | 18 | 27 | 36 | 18 |  |
| Check23 |  |  |  |  | 0.889275 |
| Pass | 1 | 40 | 13 | 5 |  |
| % | 2 | 68 | 22 | 8 |  |
| Fail/Possible fail | 0 | 7 | 5 | 0 |  |
| % | 0 | 58 | 42 | 0 |  |
| Check24 |  |  |  |  | 0.025713 |
| Pass | 0 | 22 | 6 | 2 |  |
| % | 0 | 73 | 20 | 7 |  |
| Fail/Possible fail | 0 | 0 | 3 | 0 |  |
| % | 0 | 0 | 100 | 0 |  |
| Check25 |  |  |  |  | 0.415731 |
| Pass | 0 | 29 | 8 | 2 |  |
| % | 0 | 74 | 21 | 5 |  |
| Fail/Possible fail | 0 | 4 | 4 | 1 |  |
| % | 0 | 44 | 44 | 11 |  |
| Check26 |  |  |  |  | 0.670283 |
| Pass | 1 | 20 | 7 | 0 |  |
| % | 4 | 71 | 25 | 0 |  |
| Fail/Possible fail | 0 | 1 | 1 | 0 |  |
| % | 0 | 50 | 50 | 0 |  |
| Check27 |  |  |  |  | 0.41095 |
| Pass | 0 | 28 | 6 | 1 |  |
| % | 0 | 80 | 17 | 3 |  |
| Fail/Possible fail | 0 | 3 | 2 | 1 |  |
| % | 0 | 50 | 33 | 17 |  |
| Check28 |  |  |  |  | 0.064978 |
| Pass | 1 | 44 | 16 | 2 |  |
| % | 2 | 70 | 25 | 3 |  |
| Fail/Possible fail | 3 | 14 | 8 | 4 |  |
| % | 10 | 48 | 28 | 14 |  |
| Check29 |  |  |  |  | NA |
| Pass | 0 | 14 | 0 | 0 |  |
| % | 0 | 100 | 0 | 0 |  |
| Fail/Possible fail | 0 | 1 | 0 | 0 |  |
| % | 0 | 100 | 0 | 0 |  |
| Check30 |  |  |  |  | 0.176143 |
| Pass | 0 | 20 | 5 | 3 |  |
| % | 0 | 71 | 18 | 11 |  |
| Fail/Possible fail | 4 | 40 | 19 | 3 |  |
| % | 6 | 61 | 29 | 5 |  |
| Check31 |  |  |  |  | 0.320196 |
| Pass | 0 | 18 | 4 | 1 |  |
| % | 0 | 78 | 17 | 4 |  |
| Fail/Possible fail | 3 | 34 | 12 | 5 |  |
| % | 6 | 63 | 22 | 9 |  |
| Check32 |  |  |  |  | 1.86E-07 |
| Pass | 0 | 23 | 7 | 0 |  |
| % | 0 | 77 | 23 | 0 |  |
| Fail/Possible fail | 2 | 0 | 0 | 0 |  |
| % | 100 | 0 | 0 | 0 |  |
| Check33 |  |  |  |  | 0.016509 |
| Pass | 0 | 15 | 2 | 0 |  |
| % | 0 | 88 | 12 | 0 |  |
| Fail/Possible fail | 2 | 1 | 2 | 0 |  |
| % | 40 | 20 | 40 | 0 |  |
| Check34 |  |  |  |  | 0.448697 |
| Pass | 2 | 12 | 3 | 0 |  |
| % | 12 | 71 | 18 | 0 |  |
| Fail/Possible fail | 0 | 1 | 2 | 0 |  |
| % | 0 | 33 | 67 | 0 |  |
| Check35 |  |  |  |  | 0.025306 |
| Pass | 0 | 5 | 3 | 0 |  |
| % | 0 | 63 | 38 | 0 |  |
| Fail/Possible fail | 2 | 0 | 0 | 0 |  |
| % | 100 | 0 | 0 | 0 |  |
| Check36 |  |  |  |  | 0.01612 |
| Pass | 0 | 11 | 2 | 0 |  |
| % | 0 | 85 | 15 | 0 |  |
| Fail/Possible fail | 2 | 1 | 0 | 0 |  |
| % | 67 | 33 | 0 | 0 |  |
| Check37 |  |  |  |  | 0.003021 |
| Pass | 2 | 44 | 14 | 0 |  |
| % | 3 | 73 | 23 | 0 |  |
| Fail/Possible fail | 1 | 13 | 10 | 6 |  |
| % | 3 | 43 | 33 | 20 |  |
| Check38 |  |  |  |  | 0.004087 |
| Pass | 0 | 8 | 0 | 0 |  |
| % | 0 | 100 | 0 | 0 |  |
| Fail/Possible fail | 0 | 1 | 0 | 0 |  |
| % | 0 | 100 | 0 | 0 |  |
| Check39 |  |  |  |  | 0.758334 |
| Pass | 4 | 45 | 20 | 3 |  |
| % | 6 | 63 | 28 | 4 |  |
| Fail/Possible fail | 0 | 6 | 3 | 1 |  |
| % | 0 | 60 | 30 | 10 |  |
| Check40 |  |  |  |  | NA |
| Pass | 0 | 1 | 0 | 0 |  |
| % | 0 | 100 | 0 | 0 |  |
| Check41 |  |  |  |  | 0.954479 |
| Pass | 0 | 19 | 5 | 1 |  |
| % | 0 | 76 | 20 | 4 |  |
| Fail/Possible fail | 0 | 7 | 1 | 1 |  |
| % | 0 | 78 | 11 | 11 |  |
| Check42 |  |  |  |  | 0.849334 |
| Pass | 2 | 15 | 5 | 0 |  |
| % | 9 | 68 | 23 | 0 |  |
| Fail/Possible fail | 0 | 1 | 1 | 0 |  |
| % | 0 | 50 | 50 | 0 |  |
| Check43 |  |  |  |  | 1.19E-05 |
| Pass | 3 | 53 | 20 | 3 |  |
| % | 4 | 67 | 25 | 4 |  |
| Fail/Possible fail | 1 | 2 | 2 | 0 |  |
| % | 20 | 40 | 40 | 0 |  |
| Check44 |  |  |  |  | 0.009213 |
| Pass | 1 | 53 | 12 | 3 |  |
| % | 1 | 77 | 17 | 4 |  |
| Fail/Possible fail | 2 | 2 | 3 | 0 |  |
| % | 29 | 29 | 43 | 0 |  |
| Check46 |  |  |  |  | 0.010492 |
| Pass | 3 | 56 | 15 | 4 |  |
| % | 4 | 72 | 19 | 5 |  |
| Fail/Possible fail | 1 | 4 | 9 | 2 |  |
| % | 6 | 25 | 56 | 13 |  |
| Check48 |  |  |  |  | 0.307784 |
| Pass | 0 | 20 | 3 | 1 |  |
| % | 0 | 83 | 13 | 4 |  |
| Fail/Possible fail | 0 | 4 | 3 | 0 |  |
| % | 0 | 57 | 43 | 0 |  |
| Check49 |  |  |  |  | 0.061558 |
| Pass | 2 | 41 | 7 | 5 |  |
| % | 4 | 75 | 13 | 9 |  |
| Fail/Possible fail | 2 | 19 | 16 | 1 |  |
| % | 5 | 50 | 42 | 3 |  |
| Check50 |  |  |  |  | 0.007256 |
| Pass | 2 | 38 | 9 | 2 |  |
| % | 4 | 75 | 18 | 4 |  |
| Fail/Possible fail | 1 | 1 | 5 | 0 |  |
| % | 14 | 14 | 71 | 0 |  |
| Check51 |  |  |  |  | 0.009307 |
| Pass | 1 | 46 | 9 | 5 |  |
| % | 2 | 75 | 15 | 8 |  |
| Fail/Possible fail | 0 | 2 | 6 | 1 |  |
| % | 0 | 22 | 67 | 11 |  |
| Check52 |  |  |  |  | 0.000534 |
| Pass | 0 | 52 | 15 | 3 |  |
| % | 0 | 74 | 21 | 4 |  |
| Fail/Possible fail | 0 | 0 | 2 | 3 |  |
| % | 0 | 0 | 40 | 60 |  |
| Check53 |  |  |  |  | 4.01E-05 |
| Pass | 2 | 58 | 16 | 3 |  |
| % | 3 | 73 | 20 | 4 |  |
| Fail/Possible fail | 0 | 0 | 2 | 3 |  |
| % | 0 | 0 | 40 | 60 |  |
| Check54 |  |  |  |  | 0.125697 |
| Pass | 1 | 42 | 8 | 4 |  |
| % | 2 | 76 | 15 | 7 |  |
| Fail/Possible fail | 0 | 5 | 5 | 0 |  |
| % | 0 | 50 | 50 | 0 |  |
| Check55 |  |  |  |  | 0.020699 |
| Pass | 1 | 46 | 17 | 2 |  |
| % | 2 | 70 | 26 | 3 |  |
| Fail/Possible fail | 3 | 11 | 6 | 4 |  |
| % | 13 | 46 | 25 | 17 |  |
| Check56 |  |  |  |  | 0.001213 |
| Pass | 2 | 51 | 15 | 2 |  |
| % | 3 | 73 | 21 | 3 |  |
| Fail/Possible fail | 0 | 1 | 6 | 4 |  |
| % | 0 | 9 | 55 | 36 |  |
| Check57 |  |  |  |  | 0.024062 |
| Pass | 2 | 59 | 19 | 4 |  |
| % | 2 | 70 | 23 | 5 |  |
| Fail/Possible fail | 0 | 1 | 2 | 2 |  |
| % | 0 | 20 | 40 | 40 |  |
| Check58 |  |  |  |  | 0.578957 |
| Pass | 4 | 58 | 23 | 6 |  |
| % | 4 | 64 | 25 | 7 |  |
| Fail/Possible fail | 0 | 0 | 1 | 0 |  |
| % | 0 | 0 | 100 | 0 |  |
| Check59 |  |  |  |  | 0.04412 |
| Pass | 2 | 52 | 19 | 3 |  |
| % | 3 | 68 | 25 | 4 |  |
| Fail/Possible fail | 2 | 6 | 5 | 3 |  |
| % | 13 | 38 | 31 | 19 |  |
| Check61 |  |  |  |  | 0.213261 |
| Pass | 0 | 7 | 1 | 0 |  |
| % | 0 | 88 | 13 | 0 |  |
| Fail/Possible fail | 4 | 49 | 18 | 5 |  |
| % | 5 | 64 | 24 | 7 |  |
| Check62 |  |  |  |  | NA |
| Pass | 0 | 2 | 0 | 0 |  |
| % |  |  |  |  |  |
| Check63 |  |  |  |  | 0.115586 |
| Pass | 0 | 13 | 0 | 0 |  |
| % | 0 | 100 | 0 | 0 |  |
| Fail/Possible fail | 0 | 19 | 5 | 2 |  |
| % | 0 | 73 | 19 | 8 |  |
| Check64 |  |  |  |  | 0.004098 |
| Pass | 0 | 27 | 3 | 0 |  |
| % | 0 | 90 | 10 | 0 |  |
| Fail/Possible fail | 4 | 26 | 17 | 5 |  |
| % | 8 | 50 | 33 | 10 |  |
| Check65 |  |  |  |  | 0.030546 |
| Pass | 0 | 29 | 5 | 0 |  |
| % | 0 | 85 | 15 | 0 |  |
| Fail/Possible fail | 0 | 5 | 3 | 2 |  |
| % | 0 | 50 | 30 | 20 |  |
| Check68 |  |  |  |  | 0.93847 |
| Pass | 1 | 27 | 6 | 3 |  |
| % | 3 | 73 | 16 | 8 |  |
| Fail/Possible fail | 0 | 9 | 4 | 2 |  |
| % | 0 | 60 | 27 | 13 |  |
| Check69 |  |  |  |  | 0.000341 |
| Pass | 0 | 20 | 3 | 0 |  |
| % | 0 | 87 | 13 | 0 |  |
| Fail/Possible fail | 0 | 0 | 0 | 1 |  |
| % | 0 | 0 | 0 | 100 |  |
| Check70 |  |  |  |  | 0.072207 |
| Pass | 4 | 58 | 21 | 4 |  |
| % | 5 | 67 | 24 | 5 |  |
| Fail/Possible fail | 0 | 2 | 3 | 2 |  |
| % | 0 | 29 | 43 | 29 |  |
| Check71 |  |  |  |  | 0.058104 |
| Pass | 4 | 58 | 20 | 4 |  |
| % | 5 | 67 | 23 | 5 |  |
| Fail/Possible fail | 0 | 0 | 1 | 1 |  |
| % | 0 | 0 | 50 | 50 |  |
| Check72 |  |  |  |  | NA |
| Pass | 0 | 23 | 10 | 3 |  |
| % | 0 | 64 | 28 | 8 |  |
| Check73 |  |  |  |  | 7.35E-06 |
| Pass | 0 | 47 | 13 | 3 |  |
| % | 0 | 75 | 21 | 5 |  |
| Fail/Possible fail | 2 | 0 | 1 | 1 |  |
| % | 50 | 0 | 25 | 25 |  |
| Check74 |  |  |  |  | NA |
| Pass | 1 | 53 | 22 | 4 |  |
| % | 1 | 66 | 28 | 5 |  |
| Check75 |  |  |  |  | 0.185483 |
| Pass | 4 | 56 | 18 | 5 |  |
| % | 5 | 67 | 22 | 6 |  |
| Fail/Possible fail | 0 | 1 | 3 | 1 |  |
| % | 0 | 20 | 60 | 20 |  |
| Check76 |  |  |  |  | 6.29E-06 |
| Pass | 1 | 54 | 16 | 4 |  |
| % | 1 | 72 | 21 | 5 |  |
| Fail/Possible fail | 2 | 0 | 1 | 0 |  |
| % | 67 | 0 | 33 | 0 |  |

S Table 8: Crosstabulation of response to each check and overall assessment of the trial.

Restricted to trials where the check was feasible. P-value is from a chi-squared test, corrected for small expected cell frequencies.

This analysis was not specified in the protocol.

**Section D – Impact of removing RCTs flagged by each check from meta-analyses**

| Check number | Proportion with no RCTs remaining | Mean ROR | Mean RSE | Proportion with change of significance | Mean Ratio of Confidence Intervals | Mean change in tau (heterogeneity) |
| --- | --- | --- | --- | --- | --- | --- |
| check1 | 0.076923 | 1.067575 | 1.033838 | 0 | 1.19175 | -0.043479 |
| check2 | 0.076923 | 1.01565 | 1.096208 | 0 | 1.361687 | -0.00691 |
| check3 | 0 | 1 | 1 | 0 | 1 | 0 |
| check4 | 0.038462 | 1.048155 | 1.033708 | 0.04 | 1.208692 | -0.046077 |
| check5 | 0 | 1.055945 | 1.007218 | 0.038462 | 1.072946 | -0.044305 |
| check6 | 0 | 1.036199 | 1.044269 | 0.038462 | 1.179063 | -0.018946 |
| check7 | 0 | 1.045172 | 1.029371 | 0 | 1.109751 | 0 |
| check8 | 0 | 1.049652 | 1.046225 | 0.038462 | 1.204208 | -0.016899 |
| check9 | 0.038462 | 0.994926 | 1.115166 | 0 | 1.541951 | 0.0208101 |
| check10 | 0.076923 | 1.022534 | 1.073416 | 0 | 2.92955 | -0.041087 |
| check11 | 0.076923 | 1.14336 | 1.166619 | 0.041667 | 3.299143 | -0.077773 |
| check12 | 0.076923 | 1.038257 | 1.086662 | 0.041667 | 1.306917 | -0.009139 |
| check13 | 0 | 1.091128 | 1.050305 | 0.076923 | 1.243703 | -0.010551 |
| check14 | 0.038462 | 0.945192 | 1.02746 | 0 | 1.142681 | -0.038102 |
| check15 | 0 | 0.964499 | 1.057767 | 0 | 1.283861 | 0.0200097 |
| check16 | 0.038462 | 1.026824 | 1.040405 | 0 | 1.342076 | 0 |
| check17 | 0 | 0.986989 | 1.064481 | 0 | 2.58969 | 0.0545052 |
| check18 | 0 | 1 | 1 | 0 | 1 | 0 |
| check19 | 0.038462 | 1.00821 | 1.087738 | 0 | 1.407792 | 0.0208101 |
| check20 | 0 | 1.086915 | 1.07807 | 0 | 1.280545 | 0 |
| check21 | 0.115385 | 1.041997 | 1.048509 | 0 | 1.145436 | 0 |
| check22 | 0.076923 | 1.072869 | 1.014992 | 0 | 1.101973 | -0.028038 |
| check23 | 0 | 0.989003 | 1.076584 | 0 | 1.553449 | 0.0374532 |
| check24 | 0 | 0.98824 | 1.018096 | 0 | 1.031544 | 0 |
| check25 | 0.038462 | 0.991588 | 1.013707 | 0 | 1.019258 | 0 |
| check26 | 0 | 0.977476 | 1.098852 | 0 | 4.303911 | 0.0555764 |
| check27 | 0 | 1.009158 | 1.01671 | 0 | 1.038071 | 0 |
| check28 | 0.307692 | 1.042573 | 1.126994 | 0 | 1.563817 | -0.010643 |
| check29 | 0 | 0.998316 | 1.007553 | 0 | 1.015636 | 0.0034979 |
| check30 | 0.576923 | 1.072926 | 1.327425 | 0.181818 | 2.432466 | -0.076465 |
| check31 | 0.538462 | 1.046348 | 1.105461 | 0 | 1.188237 | -0.027607 |
| check32 | 0.038462 | 1 | 1 | 0 | 1 | 0 |
| check33 | 0.038462 | 0.995323 | 1.032179 | 0 | 1.139745 | 0 |
| check34 | 0 | 1.005146 | 1.005748 | 0 | 1.00665 | 0 |
| check35 | 0.038462 | 1 | 1 | 0 | 1 | 0 |
| check36 | 0.038462 | 1 | 1 | 0 | 1 | 0 |
| check37 | 0.269231 | 1.017538 | 1.036206 | 0 | 1.190736 | -0.041406 |
| check38 | 0 | 1 | 1 | 0 | 1 | 0 |
| check39 | 0.038462 | 0.992742 | 1.030147 | 0 | 1.023254 | 0 |
| check40 | 0 | 1 | 1 | 0 | 1 | 0 |
| check41 | 0.115385 | 1 | 1 | 0 | 1 | 0 |
| check42 | 0 | 1 | 1 | 0 | 1 | 0 |
| check43 | 0 | 0.984971 | 1.042103 | 0 | 1.197444 | 0.0263879 |
| check44 | 0.038462 | 0.989824 | 1.082407 | 0 | 1.599808 | 0.0339643 |
| check45 | NA | NA | NA | NA | NA | NA |
| check46 | 0.115385 | 1.006637 | 0.966534 | 0 | 0.950602 | -0.067416 |
| check47 | NA | NA | NA | NA | NA | NA |
| check48 | 0.038462 | 1.012559 | 1.038758 | 0 | 1.396172 | -0.032923 |
| check49 | 0.269231 | 0.9519 | 1.06011 | 0 | 1.195318 | -0.060627 |
| check50 | 0.076923 | 0.975599 | 1.107089 | 0 | 4.579237 | 0.0602078 |
| check51 | 0.038462 | 0.983874 | 1.041164 | 0 | 1.199028 | 0.0274434 |
| check52 | 0 | 1.019713 | 1.04967 | 0 | 1.213673 | -0.006378 |
| check53 | 0.038462 | 1.045131 | 1.02766 | 0 | 1.112577 | 0 |
| check54 | 0.076923 | 0.958428 | 1.032632 | 0 | 1.194748 | -0.007982 |
| check55 | 0.192308 | 1.099193 | 1.080084 | 0.047619 | 1.651824 | -0.037306 |
| check56 | 0.076923 | 1.021452 | 1.085969 | 0 | 1.344648 | 0.0285869 |
| check57 | 0.038462 | 1.029005 | 1.068824 | 0 | 1.311605 | 0.0274434 |
| check58 | 0 | 1 | 1 | 0 | 1 | 0 |
| check59 | 0.115385 | 1.034538 | 1.038921 | 0 | 1.169079 | 0 |
| check60 | NA | NA | NA | NA | NA | NA |
| check61 | 0.730769 | 1.157955 | 1.102861 | 0 | 2.16515 | -0.096131 |
| check62 | 0 | 1 | 1 | 0 | 1 | 0 |
| check63 | 0.230769 | 0.970719 | 1.128507 | 0 | 5.295084 | 0.0722493 |
| check64 | 0.5 | 0.965362 | 1.092787 | 0 | 2.923555 | 0.0150294 |
| check65 | 0.076923 | 1.066875 | 1.029617 | 0.041667 | 1.30718 | -0.014333 |
| check66 | NA | NA | NA | NA | NA | NA |
| check67 | NA | NA | NA | NA | NA | NA |
| check68 | 0.076923 | 0.953858 | 1.007456 | 0 | 1.247942 | -0.036569 |
| check69 | 0 | 1 | 1 | 0 | 1 | 0 |
| check70 | 0 | 0.966134 | 1.029387 | 0 | 1.138115 | -0.006378 |
| check71 | 0 | 1.010164 | 1.012314 | 0 | 1.021717 | 0.0073661 |
| check72 | 0 | 1 | 1 | 0 | 1 | 0 |
| check73 | 0.038462 | 1 | 1 | 0 | 1 | 0 |
| check74 | 0 | 1 | 1 | 0 | 1 | 0 |
| check75 | 0 | 1.009269 | 1.011293 | 0 | 1.108798 | -0.040134 |
| check76 | 0.038462 | 0.983874 | 1.041164 | 0 | 1.199028 | 0.0274434 |

**S Table 9: Impact of removing RCTs flagged by each check (possible fail or fail) from meta-analyses with binary outcomes.**

Apart from column 1, metrics were calculated in meta-analyses with at least one trial remaining after removing flagged trials.

ROR = ratio of odds ratios, RSE – ratio of standard errors

| Check Number | Proportion with no RCTs remaining | Mean difference in SMDs | Mean RSE | Proportion with change of significance | Mean Ratio of Confidence Intervals | Mean change in tau (heterogeneity) |
| --- | --- | --- | --- | --- | --- | --- |
| check1 | 0.083333 | 0 | 1 | 0 | 1 | 0 |
| check2 | 0.125 | 0.013915 | 0.976374 | 0.047619 | 0.976374 | -0.01837 |
| check3 | 0.041667 | -0.0135 | 1.003892 | 0 | 1.003892 | -0.01677 |
| check4 | 0 | 0 | 1 | 0 | 1 | 0 |
| check5 | 0.041667 | 0 | 1 | 0 | 1 | 0 |
| check6 | 0.083333 | -0.00902 | 0.996058 | 0.045455 | 0.996058 | -0.01753 |
| check7 | 0.083333 | 0.010078 | 1.007771 | 0.045455 | 1.007771 | -0.01324 |
| check8 | 0.041667 | 0.010616 | 1.002099 | 0.043478 | 1.002099 | -0.01503 |
| check9 | 0.166667 | 0 | 1 | 0 | 1 | 0 |
| check10 | 0.041667 | -0.01819 | 0.990549 | 0 | 0.990549 | -0.01473 |
| check11 | 0.125 | -0.00846 | 1.039359 | 0 | 1.039359 | 0 |
| check12 | 0.208333 | 8.80E-05 | 1.016348 | 0.052632 | 1.016348 | -0.0203 |
| check13 | 0.083333 | 0.005596 | 1.028754 | 0.045455 | 1.028754 | 0 |
| check14 | 0.083333 | 0.005702 | 1.030208 | 0.090909 | 1.030208 | 0.005931 |
| check15 | 0.083333 | 0.012941 | 0.984567 | 0.045455 | 0.984567 | -0.01324 |
| check16 | 0 | 0 | 1 | 0 | 1 | 0 |
| check17 | 0 | 0 | 1 | 0 | 1 | 0 |
| check18 | 0.041667 | 0.012705 | 0.978429 | 0.043478 | 0.978429 | -0.01677 |
| check19 | 0.041667 | 0 | 1 | 0 | 1 | 0 |
| check20 | 0.166667 | -0.00315 | 1.025525 | 0 | 1.025525 | 0 |
| check21 | 0.166667 | 0.011461 | 1.000718 | 0.05 | 1.000718 | -0.01929 |
| check22 | 0 | 0.004418 | 0.984239 | 0 | 0.984239 | -0.01484 |
| check23 | 0.208333 | 0.002306 | 1.016085 | 0.052632 | 1.016085 | 0.004968 |
| check24 | 0 | 0 | 1 | 0 | 1 | 0 |
| check25 | 0.125 | 0.004612 | 1.036242 | 0.047619 | 1.036242 | 0.009019 |
| check26 | 0 | 0 | 1 | 0 | 1 | 0 |
| check27 | 0 | 0.018408 | 1.027713 | 0.041667 | 1.027713 | -0.00975 |
| check28 | 0.166667 | -0.01462 | 1.078594 | 0.05 | 1.078594 | -0.01546 |
| check29 | 0 | 0 | 1 | 0 | 1 | 0 |
| check30 | 0.541667 | -0.0275 | 1.013972 | 0.090909 | 1.013972 | -0.03507 |
| check31 | 0.5 | 0.004278 | 1.012417 | 0.083333 | 1.012417 | 0 |
| check32 | 0 | 0 | 1 | 0 | 1 | 0 |
| check33 | 0 | 0.002048 | 1.013928 | 0.041667 | 1.013928 | 0 |
| check34 | 0 | -0.00447 | 1.030972 | 0 | 1.030972 | 0.002888 |
| check35 | 0 | 0 | 1 | 0 | 1 | 0 |
| check36 | 0 | 0.002048 | 1.013928 | 0.041667 | 1.013928 | 0 |
| check37 | 0.291667 | 0.004411 | 1.010735 | 0 | 1.010735 | -0.0114 |
| check38 | 0.041667 | 0 | 1 | 0 | 1 | 0 |
| check39 | 0.083333 | 0.009711 | 1.024896 | 0.045455 | 1.024896 | 0 |
| check40 | 0 | 0 | 1 | 0 | 1 | 0 |
| check41 | 0.041667 | 0.0085 | 0.991891 | 0.043478 | 0.991891 | -0.0128 |
| check42 | 0.083333 | 0 | 1 | 0 | 1 | 0 |
| check43 | 0.041667 | 0.007569 | 1.042718 | 0 | 1.042718 | -0.01841 |
| check44 | 0.125 | 0 | 1 | 0 | 1 | 0 |
| check45 | NA | NA | NA | NA | NA | NA |
| check46 | 0.166667 | -0.00268 | 1.012873 | 0.05 | 1.012873 | -0.01273 |
| check47 | NA | NA | NA | NA | NA | NA |
| check48 | 0.041667 | 0 | 1 | 0 | 1 | 0 |
| check49 | 0.333333 | -0.02157 | 1.009152 | 0.0625 | 1.009152 | -0.04637 |
| check50 | 0.041667 | 0 | 1 | 0 | 1 | 0 |
| check51 | 0.125 | 0.029634 | 0.985978 | 0.047619 | 0.985978 | -0.03853 |
| check52 | 0.041667 | 0 | 1 | 0 | 1 | 0 |
| check53 | 0.041667 | 0 | 1 | 0 | 1 | 0 |
| check54 | 0.041667 | 0.010777 | 0.988552 | 0.043478 | 0.988552 | -0.01376 |
| check55 | 0.166667 | -0.01493 | 1.068346 | 0 | 1.068346 | 0.002343 |
| check56 | 0.041667 | 0.014997 | 0.995828 | 0 | 0.995828 | -0.01841 |
| check57 | 0 | -0.00185 | 1.009701 | 0 | 1.009701 | 0.002888 |
| check58 | 0.041667 | 0 | 1 | 0 | 1 | 0 |
| check59 | 0.166667 | 0 | 1 | 0 | 1 | 0 |
| check60 | NA | NA | NA | NA | NA | NA |
| check61 | 0.666667 | 0 | 1 | 0 | 1 | 0 |
| check62 | 0 | 0 | 1 | 0 | 1 | 0 |
| check63 | 0.083333 | -0.07454 | 1.462698 | 0.045455 | 1.462698 | 0.106631 |
| check64 | 0.416667 | -0.00401 | 1.028467 | 0 | 1.028467 | -0.02544 |
| check65 | 0.083333 | -0.0078 | 1.019578 | 0 | 1.019578 | 0.00315 |
| check66 | NA | NA | NA | NA | NA | NA |
| check67 | NA | NA | NA | NA | NA | NA |
| check68 | 0.083333 | -0.17304 | 1.234088 | 0.045455 | 1.234088 | -0.01619 |
| check69 | 0.041667 | 0 | 1 | 0 | 1 | 0 |
| check70 | 0.083333 | -0.0032 | 1.030323 | 0 | 1.030323 | 0.00429 |
| check71 | 0 | 0 | 1 | 0 | 1 | 0 |
| check72 | 0 | 0 | 1 | 0 | 1 | 0 |
| check73 | 0.083333 | 0 | 1 | 0 | 1 | 0 |
| check74 | 0 | 0 | 1 | 0 | 1 | 0 |
| check75 | 0 | -0.00715 | 1.017946 | 0 | 1.017946 | 0.002888 |
| check76 | 0 | 0 | 1 | 0 | 1 | 0 |

**S Table 10: Impact of removing RCTs flagged by each check (possible fail or fail) from meta-analyses with continuous outcomes.**
